# Exploring trade-offs in diagnostic algorithm, population coverage, and duration of community screening for tuberculosis

**DOI:** 10.1101/2025.08.22.25334237

**Authors:** Katherine C. Horton, Alexandra S. Richards, Alvaro Schwalb, Rein M. G. J. Houben

**Affiliations:** Department of Infectious Disease Epidemiology, London School of Hygiene and Tropical Medicine, London, UK; TB Modelling Group, London School of Hygiene and Tropical Medicine, London, UK; School of Health and Wellbeing, University of Glasgow, Glasgow, UK; Instituto de Medicina Tropical Alexander von Humboldt, Universidad Peruana Cayetano Heredia, Lima, Peru

## Abstract

**Background:** Current tuberculosis (TB) prevention and care strategies have failed to reduce disease burden at the pace required to meet global targets. Community screening may enable more rapid declines in TB burden, but evidence is limited. We used mathematical modelling to evaluate trade-offs in diagnostic algorithm, population coverage, and duration of screening.

**Methods and Findings:** We used a TB model, which recognised symptomatic and asymptomatic infectious TB (defined by whether an individual reported symptoms at screening), as well as non-infectious TB. We simulated diagnostic algorithms targeting symptomatic infectious TB (prolonged cough with confirmatory Xpert Ultra), infectious TB (Xpert Ultra), or all TB (chest X-ray), and we varied population coverage and duration. Main outcomes were estimated reduction in symptomatic TB incidence and TB mortality over a 10-year horizon.

Maximum coverage (100%) and duration (five rounds) was projected to reduce symptomatic TB incidence by 26.9% (22.8-31.5%) with the algorithm targeting symptomatic TB and 74.0% (68.5-79.1%) with the algorithm targeting infectious TB. However, incidence rebounded at the end of screening, erasing 9.8% and 15.9%, respectively, of those reductions within five years. The algorithm targeting all TB showed higher potential for rapid reductions – over 98% – with negligible rebound; however, low diagnostic accuracy of current tools led to prohibitive overdiagnosis, with 7.2 false positives per true positive in a single round of screening targeting all TB. Screening algorithms targeting broader disease definitions generally achieved greater impact with lower population coverage and/or duration. Findings were broadly similar for mortality.

**Conclusions:** We show substantial reductions in TB morbidity and mortality can be achieved by community screening, highlighting the importance of symptom-agnostic algorithms and the need to balance population coverage and duration. To maximise and sustain epidemiological impact, diagnostic tools and treatment regimens for individuals with non-infectious TB are needed.

## Introduction

Current tuberculosis (TB) prevention and care strategies have failed to reduce disease burden at the pace required to meet global targets. Whilst TB incidence and mortality have fallen 13% and 35%, respectively, over the past decade, progress remains far from the ambitious targets of the EndTB Strategy, which aims for 90% and 95% reductions, respectively, by 2035 [1]. Instead, an estimated 10.7 million people developed TB in 2024, and TB remained the leading infectious cause of mortality worldwide [3]. There is growing recognition that expanded strategies are needed to reach the millions of affected individuals and interrupt ongoing transmission [1].

Community screening offers an opportunity to reach affected individuals who are unable to access TB diagnosis and treatment through routine health services [4]. Provider-initiated diagnosis and treatment in community settings may reach individuals with limited knowledge of TB, inadequate resources to support care-seeking, or limited access to facilities that offer TB diagnosis and treatment. Community screening also presents an opportunity for diagnosis to individuals with non-infectious TB or asymptomatic infectious TB that is unlikely to prompt care-seeking [5, 6]. By extending the benefits of diagnosis and treatment to these individuals, community screening may reduce adverse health, social, and economic outcomes for affected individuals and interrupt transmission within affected communities [7].

The reach of community screening translated into rapid declines in TB burden in the twentieth century in many countries that now have a low burden of disease [8]. However, the approach was widely discontinued in the 1970s due to costs and logistics, and focus shifted towards strengthening diagnosis and treatment within health systems [9]. Renewed interest in community screening emerged in the 2000s, with the World Health Organization recommending systematic screening in certain high burden populations [4, 7], but contemporary evidence on the effectiveness of community screening is limited [10, 11]. Few randomised controlled trials evaluating the epidemiological impact of systematic screening have been conducted, and vastly different approaches were used in existing studies.

The population-level effectiveness of community screening will depend on considerations including diagnostic algorithm, population coverage, and duration of screening. While the optimal approach might combine a highly accurate algorithm targeting all individuals with TB, with high population coverage and long duration of screening, maximising all of these factors simultaneously is rarely feasible due to resource constraints. As such, decisions are required to maximise individual and population health impact whilst minimising demands on financial and human resources. Evidence to inform such decisions is limited, and the relative influence of different aspects of screening approaches is challenging to isolate and compare across randomised controlled trials and other studies.

We therefore used mathematical modelling to evaluate the potential epidemiological impact of different community screening approaches for TB, exploring trade-offs in diagnostic algorithm, population coverage, and duration of screening.

## Methods

Our analysis considers community screening approaches within a framework that acknowledges a spectrum of disease states including symptomatic infectious TB (sTB, bacteriologically confirmed with report of TB symptoms at screening), asymptomatic infectious TB (aTB, bacteriologically confirmed with no report of TB symptoms at screening), and non-infectious TB (nTB, macroscopic pathology of TB, bacteriologically unconfirmed, regardless of symptoms) [5, 6, 13]. We focus solely on pulmonary TB in adults and adolescents (age ≥ 15 years).

### Model structure and parameterisation

We expanded a deterministic compartmental cohort model of *Mycobacterium tuberculosis* (*Mtb*) infection and TB disease (sTB, aTB, and nTB), described elsewhere, to incorporate *Mtb* transmission and TB treatment (Fig 1). In our model, susceptible individuals and those who have previously cleared infection are at risk of infection, and individuals who have recovered from nTB are at risk of reinfection (with an assumption of partial protection), as are those who have been treated for sTB (with increased risk [15]). aTB is considered relatively less infectious than sTB [16].

**Fig 1:**
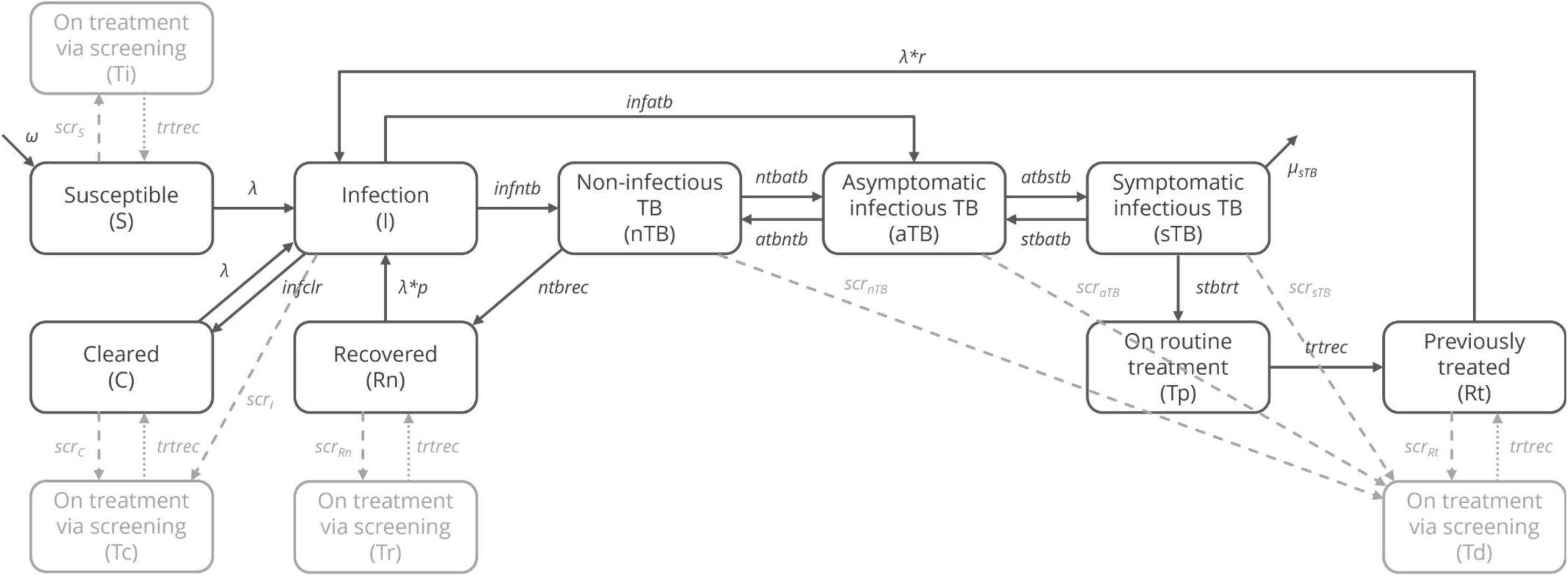
Model structure. Core model states and transitions are shown in black. All states experience background mortality (not shown). Implementation of community screening approaches is shown in grey, with dashed lines showing pathways onto treatment via screening and dotted lines showing pathways upon treatment completion for treatment via screening. Rates of transitions onto treatment via screening (indicated by “scr” with a subscript) are defined by screening algorithms.

The model was implemented with a one-month timestep for a fixed population of 100,000 individuals with susceptible individuals entering the model set equivalent to those exiting via background mortality or TB-specific mortality to maintain equilibrium. Equations for the base model without screening are provided in Model Equations in S1 Text. All transition rates were assigned non-informative uniform priors (except treatment completion and background mortality rates). Median values and ranges for prior distributions are presented in Supplemental Table 1 in S1 Text [13–19].

### Model calibration

The model was calibrated independently to three levels of infectious TB prevalence: 1,000, 500, and 250 per 100,000 population (± 10%), each after a 300-year run-in period to reach a steady state. Calibration was performed in R statistical software using history matching and emulation (hmer) [21] to identify a set of non-implausible parameter values. After achieving a set of posterior parameters consistent with the target prevalence, we used hmer emulators to sample a wider selection of non-implausible points. We then randomly selected 1,000 posterior parameter sets from those consistent with the target prevalence.

### Screening approach

We examined algorithms designed to target different presentations within the spectrum of TB based on currently available screening and diagnostic tools: sTB by symptom screening with confirmatory Xpert Ultra for those with prolonged cough (“Cough+Xpert”); infectious TB (sTB+aTB) by Xpert Ultra for all, regardless of symptoms (“Xpert”); and all TB (sTB+aTB+nTB) by chest X-ray for all, regardless of symptoms (“CXR”). The algorithm targeting all TB did not require bacteriological confirmation in order to diagnose individuals with nTB. In scenarios using the algorithm targeting infectious TB, we assumed that 100% of individuals with infectious TB were able to provide an acceptable sample for testing, while only 60% of the remaining population was able to do so. In scenarios using the algorithm targeting sTB, we assumed all those with prolonged cough were able to provide an acceptable sample for testing due to intensified efforts following report of prolonged cough. Algorithms were defined using probabilities of a positive test result, specific to each model state, which were sampled independently. Probabilities of a positive test were broadly based on sensitivity for sTB and specificity for non-TB states [4, 22–24], complemented by data from national TB prevalence surveys [25, 26], with assumptions where no data were available (Supplemental Table 3 in S1 Text and Supplemental Table 4 in S1 Text). For simplicity, we assumed 100% treatment initiation and treatment success among individuals who tested positive for TB.

Community screening was implemented in 12-month rounds. The entire population (adults and adolescents) was eligible for screening over the course of each round of screening, with one-twelfth of the population eligible for screening each month. We considered scenarios with screening coverage ranging from 0% to 100% of the eligible population, distributed proportionally across model states with no increased or decreased likelihood of undergoing screening for those with TB and/or symptoms. We varied duration of screening from one to five consecutive rounds of annual screening with constant diagnostic algorithm and population coverage over time for each scenario.

### Screening implementation

Any individual could be placed on treatment via screening except those already on treatment. Individuals with nTB, aTB, or sTB moved to the previously treated compartment upon completion of treatment, while infected individuals moved to the cleared compartment. Individuals who were susceptible, had cleared infection, or had recovered from nTB at the time they were placed on treatment via screening returned to the same respective compartment upon treatment completion.

Screening was implemented with an expanded model that tracked individuals who had been considered for screening and placed individuals who tested positive for TB during screening on treatment. This process used the deSolve event function [27], which interrupts integration routines to allow changes in model states at designated timepoints. Screening implementation is described in greater detail in Screening Implementation in S1 Text.

### Analysis

Each screening approach was implemented for 1,000 parameter sets consistent with each target prevalence (1,000, 500, and 250 per 100,000 population). We report the number of individuals with positive screening results for TB, with true positives defined as individuals with sTB, aTB, or nTB, and false positives defined as individuals in any non-TB state. We examined the impact on disease burden by estimating reductions in sTB incidence and TB mortality at the end of screening and ten years after the first year of screening (2035), as well as episodes of sTB and TB-associated deaths averted over the 10-year period from the first year of screening (2025-2035). Results are reported as median values with 95% uncertainty intervals (UIs) across parameter sets.

### Data accessibility

Replication data and analyses scripts are available on GitHub at https://github.com/ERC-TBornotTB/ScreeningTradeOffs.

## Results

### Model calibration

Median values and ranges for posterior distributions are shown in Supplemental Table 1 in S1 Text. Epidemiological burden estimates for each calibrated model are shown in Supplemental Table 2 in S1 Text.

### Diagnostic algorithm

The number of individuals with infectious TB (aTB or sTB) reached by the algorithm targeting sTB was less than half the number reached by either the algorithm targeting infectious TB (aTB+sTB) or the algorithm targeting all TB (nTB+aTB+sTB); the algorithm targeting all TB reached significantly more individuals with nTB than either of the other algorithms (Supplemental Table 5 in S1 Text). Findings were similar across coverages, durations, and baseline prevalence levels.

However, we estimated that for each individual correctly diagnosed with TB, 2.8 (1.5-4.6), 1.2 (0.9-1.6), and 7.2 (5.5-9.4) individuals without TB tested positive for algorithms targeting sTB, infectious TB, and all TB, respectively, in a single round of screening for a population with baseline prevalence of 500 per 100,000 (Fig 2). The number of false positives per true positive was lower for baseline prevalence of 1,000 per 100,000 and higher for baseline prevalence of 250 per 100,000 (Supplemental Figure 2 in S1 Text).

**Fig 2:**
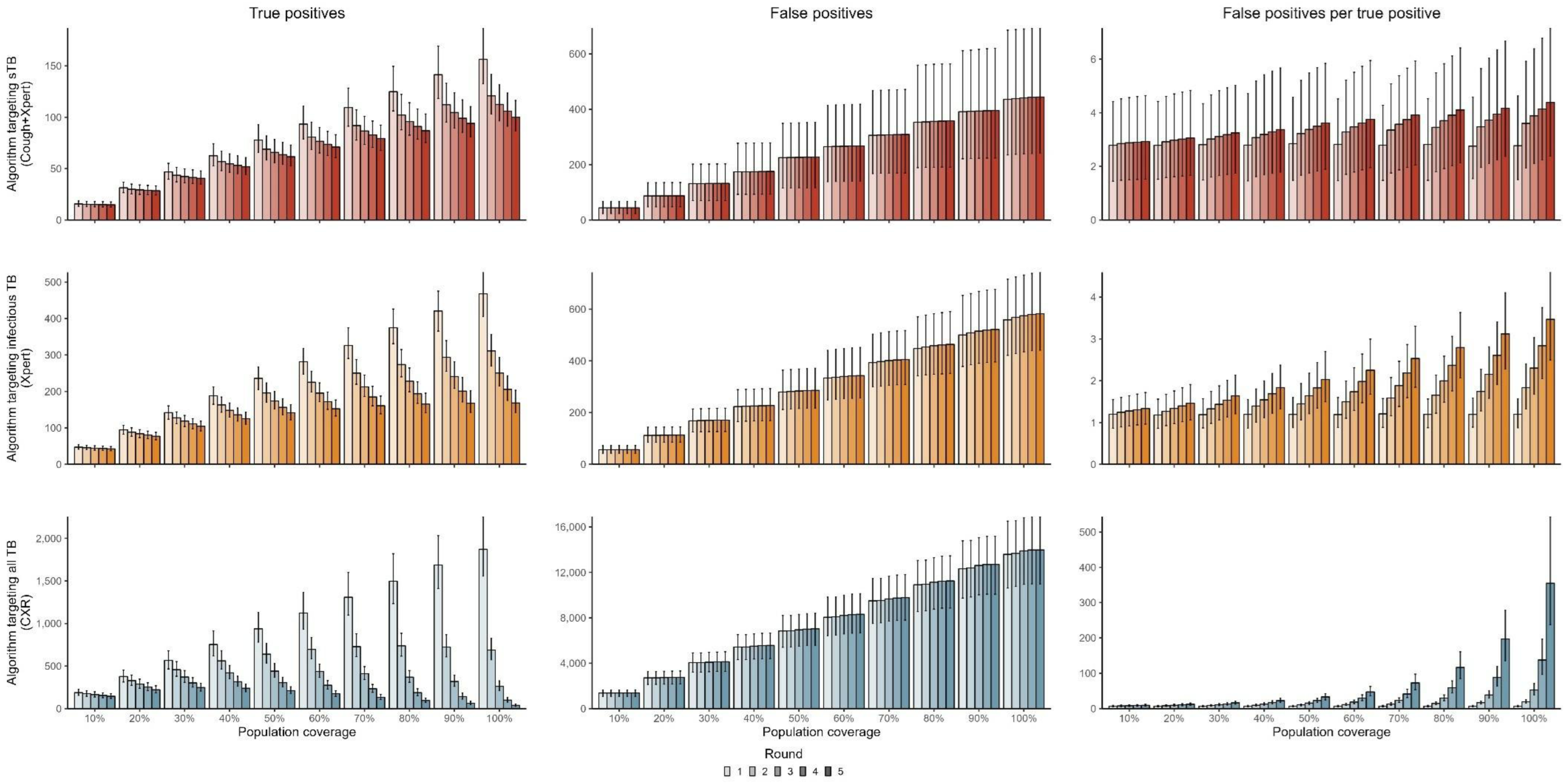
Number of true positive results, false positive results, and false positive results per true positive result for each round of screening (up to five rounds) by diagnostic algorithm and population coverage. sTB: symptomatic infectious TB; Cough+Xpert: prolonged cough with confirmatory Xpert Ultra; Xpert: Xpert Ultra; CXR: chest X-ray.

For a single round of screening, the algorithm targeting infectious TB required eight times as many confirmatory tests with Xpert Ultra tests as the algorithm targeting sTB across coverage and baseline prevalence levels (Supplemental Figure 3 in S1 Text).

### Population coverage

For a single round of screening, the number of individuals correctly diagnosed increased proportionally with population coverage across algorithms (Fig 2). While this linear association would be expected, the effort required to achieve increased coverage would be unlikely to scale linearly. Relative increases were similar for the number of individuals who received false positive results and across baseline prevalence levels.

### Duration of screening

At low population coverage (10%), similar numbers of individuals were correctly diagnosed with TB in each round of screening with the algorithm targeting sTB; true positives fell slightly with each round of screening for algorithms targeting infectious TB or all TB (Fig 2). In contrast, at maximum coverage (100%), the number of individuals correctly diagnosed in the second round was 20%, 30%, and 60% lower than the first round for the algorithm targeting sTB, the algorithm targeting infectious TB, and the algorithm targeting all TB, respectively. With each subsequent round of screening, the number of true positive results fell, with more substantial declines for algorithms targeting infectious TB or all TB, reflecting relative impact on disease burden across algorithms. The number of false positives increased slightly (<2% per round) with each subsequent round across population coverage and algorithm. The number of false positives per true positive increased rapidly as high population coverage led to declines in TB prevalence, especially for the algorithm targeting all TB (Fig 2).

### Epidemiological impact

Maximum coverage (100%) and duration (five rounds) with the algorithm targeting sTB resulted in a 26.9% (22.8-31.5%) decline in incidence at the end of screening (Fig 3), which rebounded by 9.8% (7.4-12.9%) within five years. The long-term reduction in incidence 10 years after the start of screening (23.7%, 20.1-28.0%) could also be achieved with the algorithm targeting infectious TB within three rounds with 70% coverage, four rounds with 60% coverage, or five rounds with 50% coverage, or with the algorithm targeting all TB within one round with 60% coverage, two rounds with 40% coverage, three rounds with 30% coverage, or four rounds with 20% coverage (Table 1, orange borders).

**Fig 2:**
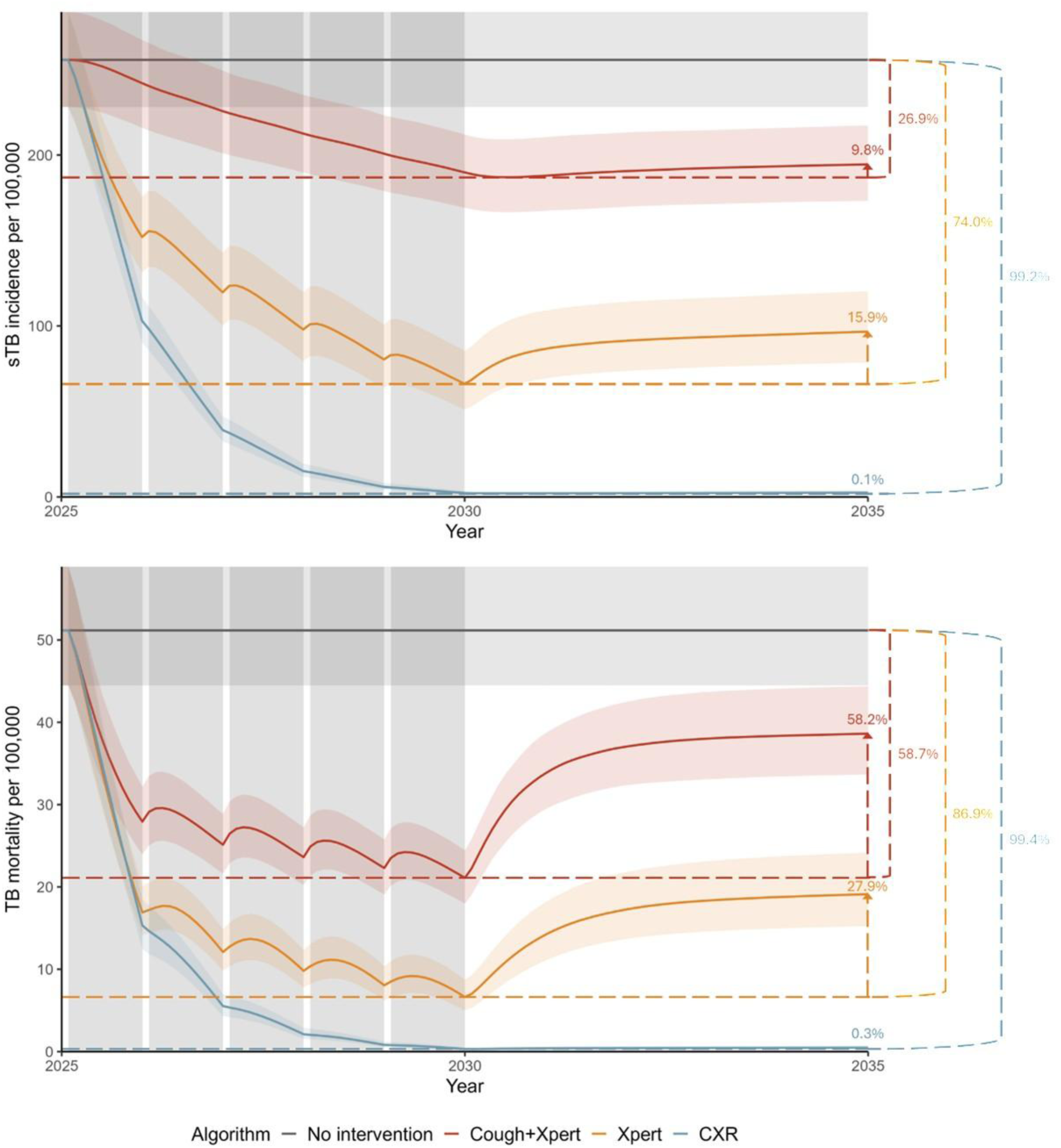
Projected incidence of symptomatic TB (sTB, top) and TB mortality (bottom) for maximum population coverage (100%) and duration of screening (5 years) for diagnostic algorithms targeting sTB (Cough+Xpert), infectious TB (Xpert), and all TB (CXR) in a population with baseline prevalence of 500 per 100,000. Horizontal dashed lines show median reduction at the end of screening for each algorithm. Vertical arrows indicate median rebound in the five years following the end of screening for each algorithm.

**Table 1:** Projected reductions in symptomatic TB incidence 10 years after the start of screening by diagnostic algorithm, population coverage, and duration in a population with baseline prevalence of 500 per 100,000 population. Orange borders show coverage and duration combinations that could achieve the same impact as maximum coverage and duration with the algorithm targeting symptomatic infectious TB; yellow show coverage and duration combinations that could achieve the same impact as maximum coverage and duration with the algorithm targeting infectious TB.

| Projected reduction in 2035 sTB incidence |  |  |  |  |  |  |  |  |  |  |
| --- | --- | --- | --- | --- | --- | --- | --- | --- | --- | --- |
| Rounds | Population coverage |  |  |  |  |  |  |  |  |  |
|  | 10% | 20% | 30% | 40% | 50% | 60% | 70% | 80% | 90% | 100% |
| Algorithm targeting symptomatic infectious TB (Cough+Xpert) |  |  |  |  |  |  |  |  |  |  |
| 1 | 0.6%<br>(0.5-0.7%) | 1.2%<br>(0.9-1.4%) | 1.7%<br>(1.4-2.1%) | 2.3%<br>(1.9-2.8%) | 2.9%<br>(2.4-3.6%) | 3.5%<br>(2.9-4.3%) | 4.1%<br>(3.4-5%) | 4.7%<br>(3.8-5.7%) | 5.3%<br>(4.3-6.4%) | 5.9%<br>(4.8-7.2%) |
| 2 | 1.2%<br>(0.9-1.4%) | 2.3%<br>(1.9-2.8%) | 3.4%<br>(2.8-4.1%) | 4.5%<br>(3.7-5.5%) | 5.6%<br>(4.7-6.7%) | 6.6%<br>(5.5-8%) | 7.6%<br>(6.3-9.1%) | 8.6%<br>(7.2-10.3%) | 9.6%<br>(7.9-11.6%) | 10.5%<br>(8.7-12.8%) |
| 3 | 1.7%<br>(1.4-2.1%) | 3.4%<br>(2.8-4.2%) | 5.1%<br>(4.2-6.1%) | 6.6%<br>(5.5-8.1%) | 8.2%<br>(6.8-9.8%) | 9.6%<br>(8-11.6%) | 11%<br>(9.2-13.2%) | 12.4%<br>(10.4-15.2%) | 13.8%<br>(11.5-16.4%) | 15%<br>(12.6-17.9%) |
| 4 | 2.3%<br>(1.9-2.8%) | 4.6%<br>(3.8-5.5%) | 6.7%<br>(5.6-8.2%) | 8.8%<br>(7.2-10.6%) | 10.8%<br>(9-12.9%) | 12.6%<br>(10.6-15.1%) | 14.4%<br>(12.2-17.2%) | 16.1%<br>(13.5-19.4%) | 17.8%<br>(15-21.3%) | 19.4%<br>(16.2-23%) |
| 5 | 2.9%<br>(2.4-3.6%) | 5.7%<br>(4.7-7%) | 8.4%<br>(6.9-10.1%) | 10.9%<br>(9.1-13%) | 13.3%<br>(11.1-15.9%) | 15.5%<br>(13.3-18.5%) | 17.7%<br>(15.1-21.1%) | 19.9%<br>(16.5-23.7%) | 21.8%<br>(18.3-26%) | 23.7%<br>(20.1-28.0%) |
| Algorithm targeting infectious TB (Xpert) |  |  |  |  |  |  |  |  |  |  |
| 1 | 1.8%<br>(1.5-2.1%) | 3.6%<br>(3-4.2%) | 5.5%<br>(4.6-6.4%) | 7.3%<br>(6.1-8.4%) | 9.2%<br>(7.8-10.6%) | 11.1%<br>(9.4-12.8%) | 12.9%<br>(11-14.9%) | 14.8%<br>(12.4-17.4%) | 16.7%<br>(14.1-19.6%) | 18.6%<br>(15.8-21.8%) |
| 2 | 3.6%<br>(3.1-4.2%) | 7.1%<br>(6.1-8.2%) | 10.6%<br>(9-12.3%) | 13.9%<br>(11.9-16.2%) | 17.2%<br>(14.8-19.9%) | 20.3%<br>(17.5-23.5%) | 23.4%<br>(20.1-27%) | 26.4%<br>(22.6-30.1%) | 29.3%<br>(25.2-33.7%) | 32%<br>(27.7-36.5%) |
| 3 | 5.4%<br>(4.6-6.3%) | 10.5%<br>(9-12.3%) | 15.5%<br>(13.3-17.9%) | 20.3%<br>(17.3-23.1%) | 24.6%<br>(21.2-28.3%) | 28.9%<br>(25-32.7%) | 32.9%<br>(28.6-37.1%) | 36.6%<br>(31.6-41.5%) | 40.2%<br>(34.6-45.5%) | 43.3%<br>(37.8-48.9%) |
| 4 | 7.2%<br>(6.1-8.3%) | 13.9%<br>(12-16%) | 20.2%<br>(17.4-23.2%) | 26.1%<br>(22.5-29.8%) | 31.5%<br>(27.2-35.8%) | 36.6%<br>(31.9-41.6%) | 41.3%<br>(36.5-46.4%) | 45.5%<br>(39.6-51%) | 49.6%<br>(44-55.2%) | 53.4%<br>(47.3-58.8%) |
| 5 | 9%<br>(7.6-10.4%) | 17.2%<br>(14.9-19.8%) | 24.8%<br>(21.3-28.5%) | 31.7%<br>(27.5-35.8%) | 37.9%<br>(33.2-42.8%) | 43.6%<br>(38.5-48.9%) | 48.7%<br>(42.8-54.5%) | 53.3%<br>(47.4-59.2%) | 57.5%<br>(51.6-63.3%) | 61.4%<br>(55.4-67.3%) |
| Algorithm targeting all TB (CXR) |  |  |  |  |  |  |  |  |  |  |
| 1 | 5.2%<br>(4.8-5.5%) | 10.5%<br>(9.8-11.2%) | 15.9%<br>(14.8-17%) | 21.4%<br>(20-22.8%) | 26.9%<br>(25-28.8%) | 32.7%<br>(30.5-34.8%) | 38.6%<br>(35.9-41%) | 44.4%<br>(41.6-47.4%) | 50.6%<br>(47.3-54%) | 56.8%<br>(53.2-60.6%) |
| 2 | 10.3%<br>(9.5-10.9%) | 20.2%<br>(18.8-21.4%) | 29.7%<br>(27.9-31.5%) | 38.8%<br>(36.4-41.2%) | 47.6%<br>(44.7-50.1%) | 55.6%<br>(52.5-58.4%) | 63.2%<br>(60.1-66.3%) | 70.3%<br>(66.8-73.3%) | 76.8%<br>(73.3-79.6%) | 82.5%<br>(79.1-85.4%) |
| 3 | 15.1%<br>(14.2-16.1%) | 29%<br>(27.3-30.8%) | 41.7%<br>(39.3-43.8%) | 52.9%<br>(50.2-55.4%) | 62.7%<br>(59.9-65.5%) | 71.4%<br>(68.5-74.1%) | 78.6%<br>(75.7-81.0%) | 84.6%<br>(81.9-86.9%) | 89.4%<br>(87-91.4%) | 93.1%<br>(91.0-94.8%) |
| 4 | 19.9%<br>(18.6-21.1%) | 37.2%<br>(35.1-39.1%) | 51.8%<br>(49.3-54.3%) | 64%<br>(61.2-66.7%) | 73.8%<br>(71.3-76.4%) | 81.7%<br>(79.1-84%) | 87.7%<br>(85.4-89.7%) | 92.2%<br>(90.3-93.7%) | 95.3%<br>(93.8-96.5%) | 97.4%<br>(96.4-98.2%) |
| 5 | 24.5%<br>(23.1-25.9%) | 44.6%<br>(42.1-46.6%) | 60.4%<br>(57.7-62.8%) | 72.7%<br>(69.8-75.2%) | 81.9%<br>(79.5-84.1%) | 88.5%<br>(86.4-90.2%) | 93.1%<br>(91.4-94.5%) | 96.1%<br>(94.9-97%) | 97.9%<br>(97.1-98.6%) | 99%<br>(98.5-99.4%) |

The algorithm targeting infectious TB, implemented with maximum coverage and duration, resulted in a 74.0% (68.5-79.1%) decline in sTB incidence at the end of screening. Five years later, incidence rebounded by 15.9% (13.5-18.6%) for a 10-year decline of 61.4% (55.4-67.3%). The algorithm targeting all TB could achieve the same long-term reduction in incidence within two rounds of screening with 90% coverage, three rounds with 60% coverage, four rounds with 50% coverage, or five rounds with 40% coverage (Table 1, yellow borders).

The algorithm targeting all TB with maximum coverage and duration was projected to reduce incidence by 99.2% (98.8-99.4%) at the end of screening, with negligible rebound (0.1%, 0.1-0.2%), which could not be achieved with other algorithm.

Across coverages and durations, the rebound in incidence at the end of screening erased 10-25% of incidence reductions for the algorithm targeting sTB, 16-55% for the algorithm targeting infectious TB, and 0-16% for the algorithm targeting all TB (Supplemental Table 16 in S1 Text). Relative epidemiological impact on incidence was similar across baseline prevalence levels.

Long-term impact on TB-associated deaths averted and TB mortality ten years after the start of screening was broadly similar to impact on sTB episodes averted and sTB incidence, respectively (Supplemental Table 12 in S1 Text and Supplemental Table 13 in S1 Text). However, the rebound in mortality at the end of screening was more extreme, erasing 58-87% of mortality reductions for the algorithm targeting sTB, 28-72% for the algorithm targeting infectious TB, and 0-25% for the algorithm targeting all TB (Supplemental Table 17 in S1 Text).

## Discussion

Community screening has the potential to substantially reduce TB morbidity and mortality when implemented with a sufficiently strong balance of diagnostic algorithm, population coverage, and duration of screening. While the same level of epidemiological impact can be achieved through multiple combinations of these factors, screening approaches that target all TB generally have higher impact with lower population coverage and/or duration, albeit with higher false positives per true positive. To maximise and sustain epidemiological impact, diagnostic tools and treatment regimens for individuals with non-infectious TB are needed.

Our work highlights the importance of symptom-agnostic algorithms, rather than symptom-based algorithms that miss affected individuals who do not report symptoms on screening. Findings from randomized controlled trials of systematic screening support the benefit of a symptom-agnostic approach. Crucially, the ACT3 trial in Viet Nam found that screening adults using GeneXpert annually over a three-year period resulted in a 44% decline in TB prevalence, whereas no impact on prevalence was found in the recent TREATS trial in Zambia and South Africa, in which adults underwent symptom screening before bacteriological testing or other trials with a symptom screen as entry point [10, 28]. Implementing symptom-agnostic screening is likely to require more human, laboratory, and financial resources than symptom-based screening. Additional efforts may also be required to avoid loss to follow-up and ensure treatment success for individuals who do not report symptoms. However, the increased population impact afforded by such efforts is key.

Our findings also highlight the potential value of developing an algorithm that can reach beyond infectious TB. We found much greater reductions in disease burden through approaches targeting all TB, including non-infectious disease, which recalls the historical success of mass radiography campaigns [8, 29]. Our results also suggest that treating this reservoir of individuals with early TB enables sustained reduction in disease burden when screening ends. However, the feasibility of treating these individuals is challenged by our limited understanding of this disease state. Current tools are not sufficiently capable of distinguishing individuals with non-infectious TB who require treatment from individuals who do not have TB, nor do we understand appropriate treatment regimens for these individuals, nor would we expect high levels of treatment uptake among individuals with no bacteriological evidence of disease. Our modelling approach used a CXR algorithm to illustrate the potential impact of screening approaches that detect individuals across all TB states. However, this standalone approach is not currently a feasible way forward due to the high number of false positives it would generate and the potential harm in placing individuals who do not have TB on treatment [30]. As such, we do not recommend initiating treatment via CXR in community screening. Instead, our results call for the development of diagnostic tools that are sensitive to and treatment regimens that are acceptable for individuals with non-infectious TB.

In our projections of sTB incidence and TB mortality during and after community screening, we observe that both incidence and mortality rebound after each round of screening with algorithms targeting sTB or infectious TB. We believe this is due to the large pool of individuals with non-infectious TB who are not detected in meaningful numbers by bacteriology-focussed approaches and remain in the population while at high risk of progression to infectious TB [31]. This hypothesis is supported by our results from scenarios using the algorithm targeting all TB, which is the only algorithm that does not rely on bacteriological confirmation. Neither incidence nor mortality rebounded meaningfully at the end of screening in these scenarios, and so reductions in burden achieved during screening were sustained afterwards. While challenging, our results highlight the potential value of identifying approaches to either treat or monitor those with radiographic evidence of TB, but negative bacteriological results. These findings could also support a need to consider alternative strategies when screening ends, such as targeted screening in high burden populations or intensified contact investigation, to ensure reductions in morbidity and mortality are sustained over time.

Our work shows the importance of population coverage and duration of screening, with limited impact attributed to single rounds of screening or to multiple rounds of screening with low population coverage, particularly for algorithms targeting only symptomatic infectious TB. It is important to note that our work assumes individuals with TB are neither more nor less likely to be screened than other individuals in the population, so population coverage here reflects an equal coverage among individuals with and without TB. If screening reaches a lower proportion of individuals with TB than the proportion of the population that is screened, impact will be reduced [32]. We also recognise that the effort required to achieve true population coverage will not scale linearly, and reaching high levels of population coverage will be difficult in many settings, highlighting the importance of balancing algorithm, coverage, and duration to achieve the desired level of impact with manageable effort.

Results presented here are intended to be illustrative. Our analysis focused on a homogeneous population with random mixing and did not incorporate geographic or individual differences in TB-associated risks, social contact patterns, or screening acceptance. Individuals who participate in community screening are unlikely to be evenly distributed across model states, and it is highly unlikely that 100% coverage could be achieved in any setting. However, as our work shows, high coverage is not necessarily needed for substantial impact. Our modelling approach recognises the spectrum of disease presentations, which required some assumptions where empirical evidence is not available, and we did not explicitly model asymptomatic, pathologically negative, infectious TB. We also did not model drug resistance and so cannot commented on the impact of community screening on drug resistance levels. Moreover, our work required assumptions about the performance of screening and diagnostic tools across model states where data are not currently available. We were ambitious in our assumptions about production of adequate samples for testing and treatment acceptance among diagnosed individuals, particularly for those without sTB; lower levels of sample provision and/or treatment success would reduce reported impacts. While we acknowledge limitations, despite attempts to use the best available data, they should not affect the qualitative observations on diagnostic algorithm, population coverage, and duration of screening.

Our use of mathematical modelling allowed us to isolate the impact of different components of community screening approaches and examine a wider array of approaches than would be feasible in randomised controlled trials or community studies. Furthermore, while most community screening programmes assess impact by measuring changes in case notifications, a modelling approach allowed us to quantify impact beyond the individuals directly reached by screening programmes and estimate changes in TB incidence and mortality across the population. Our work highlights opportunities for community screening to achieve the substantial reductions in TB morbidity and mortality required to end TB with approaches that balance diagnostic algorithm, population coverage, and duration.

## Author contributions

KCH: Conceptualisation, Methodology, Software, Formal analysis, Visualisation, Writing - Original Draft. ASR: Conceptualisation, Methodology, Software, Writing - Review & Editing. AS: Methodology, Writing - Review & Editing. RMGJH: Conceptualisation, Methodology, Visualisation, Writing - Review & Editing, Supervision, Funding acquisition.

