## Supplementary material for "Exploring trade-offs in diagnostic algorithm, population coverage, and duration of community screening for tuberculosis": S1 Text

### SUPPLEMENTAL MATERIALS

### I. Model equations

The base model without screening is described by the following set of equations:

$$N(t) = S(t) + I(t) + C(t) + Rn(t) + nTB(t) + aTB(t) + sTB(t) + Tp(t) + Rt(t)$$

$$\lambda(t) = \left(\beta * \frac{(t * aTB(t) + sTB(t))}{N(t)}\right)$$

$$\omega(t) = \mu * N(t) + \mu_{sTB} * sTB(t)$$

$$\frac{dS(t)}{dt} = -(\lambda(t) + \mu) * S(t) + \omega(t)$$

$$\frac{dC(t)}{dt} = -(\lambda(t) + \mu) * C(t) + infclr * I(t)$$

$$\frac{dRn(t)}{dt} = -(\lambda(t) * p + \mu) * Rn(t) + ntbrec * nTB(t)$$

$$\frac{dI(t)}{dt} = -(infclr + infntb + infatb + \mu) * I(t) + \lambda(t) * (S(t) + p * Rn(t) + r * Rt(t))$$

$$\frac{dnTB(t)}{dt} = -(ntbrec + ntbatb + \mu) * nTB(t) + infntb * I(t) + atbntb * aTB(t)$$

$$\frac{daTB(t)}{dt} = -(atbntb + atbstb + \mu) * aTB(t) + infatb * I(t) + ntbatb * nTB(t) + stbatb * sTB(t)$$

$$\frac{dsTB(t)}{dt} = -(stbatb + stbtrt + \mu + \mu_{sTB}) * sTB(t) + atbstb * aTB(t)$$

$$\frac{dTp(t)}{dt} = -(trtrec + \mu) * Tp(t) + stbtrt * sTB(t)$$

$$\frac{dRt(t)}{dt} = -(\lambda(t) * r + \mu) * Rt(t) + trtrec * Tp(t)$$

### II. Model calibration

Supplemental Table 1: Prior and posterior parameters for each calibration

| Parameter | Description | Prior median and range | Posterior median and range | | | Ref. |
|---|---|---|---|---|---|---|
| | | | Prevalence 1,000/100,000 | Prevalence 500/100,000 | Prevalence 250/100,000 | |
| beta | Transmission coefficient | 6 (0-12) | 2.55 (1.79-3.19) | 2.83 (2.42-3.26) | 1.61 (1.37-1.86) | - |
| p | Relative risk of infection following recovery from non-infectious TB | 0.22 (0.14-0.30) | 0.20 (0.16-0.26) | 0.22 (0.17-0.27) | 0.20 (0.17-0.23) | [1] |
| r | Relative risk of infection for previously treated individuals | 3.21 (2.14-4.27) | 2.99 (2.26-3.93) | 3.24 (2.49-3.93) | 3.09 (2.30-4.11) | [2] |
| t | Transmission from asymptomatic infectious TB relative to symptomatic infectious TB | 0.81 (0.62-1.00) | 0.78 (0.65-0.93) | 0.87 (0.75-0.96) | 0.80 (0.71-0.89) | [3] |
| infclr | Rate of clearance from infection | 2.12 (0.93-3.30) | 2.11 (1.40-2.77) | 2.57 (2.02-3.11) | 1.95 (1.65-2.35) | [4] |
| infntb | Rate of progression from infection to non-infectious TB | 0.14 (0.04-0.23) | 0.11 (0.06-0.16) | 0.09 (0.06-0.13) | 0.11 (0.08-0.13) | [4] |
| infatb | Rate of progression from infection to asymptomatic infectious TB | 0.06 (0.01-0.10) | 0.04 (0.02-0.05) | 0.03 (0.02-0.05) | 0.06 (0.04-0.08) | [4] |
| ntbrec | Rate of recovery from non-infectious TB | 0.19 (0.14-0.23) | 0.19 (0.16-0.21) | 0.18 (0.16-0.21) | 0.18 (0.16-0.20) | [4] |
| ntbatb | Rate of progression from non-infectious TB to asymptomatic infectious TB | 0.25 (0.21-0.28) | 0.25 (0.23-0.27) | 0.25 (0.22-0.27) | 0.24 (0.22-0.25) | [4] |
| atbntb | Rate of recovery from subclinical to non-infectious TB | 1.64 (1.24-2.03) | 1.64 (1.45-1.86) | 1.67 (1.45-1.89) | 1.71 (1.52-1.86) | [4] |
| atbstb | Rate of progression from asymptomatic infectious TB to symptomatic infectious TB | 0.75 (0.56-0.94) | 0.77 (0.65-0.90) | 0.75 (0.65-0.86) | 0.75 (0.64-0.87) | [4] |
| stbatb | Rate of recovery from symptomatic infectious TB to asymptomatic infectious TB | 0.59 (0.46-0.72) | 0.58 (0.49-0.68) | 0.60 (0.51-0.70) | 0.56 (0.47-0.64) | [4] |
| stbtrt | Rate of routine treatment initiation from symptomatic infectious TB | 0.67 (0.57-0.77) | 0.68 (0.60-0.74) | 0.68 (0.61-0.75) | 0.68 (0.62-0.73) | [5] |
| trtrec | Rate of treatment completion | 2.00 | 2.00 | 2.00 | 2.00 | [6] |
| $\mu_{sTB}$ | TB-specific mortality rate | 0.33 (0.28-0.38) | 0.33 (0.29-0.36) | 0.32 (0.30-0.35) | 0.32 (0.29-0.35) | [4] |
| $\mu$ | Background mortality rate | 0.0137 | 0.0137 | 0.0137 | 0.0137 | [7] |

Supplemental Table 2: Epidemiological estimates from models calibrated to infectious TB prevalence levels of 1,000, 500, and 250 per 100,000 population (± 10%). Table shows median values and 95% uncertainty intervals. infTB: infectious TB; sTB: symptomatic infectious TB; nTB: non-infectious TB.

Epidemiological estimates from calibrated models

| | Target prevalence per 100,000 | | |
|---|---|---|---|
| Estimate | 250 | 500 | 1,000 |
| infTB prevalence (per 100,000) | 249.5 (233.9-265.8) | 502.6 (467.3-535.8) | 1003.7 (931.9-1070.7) |
| sTB incidence (per 100,000) | 126.9 (111.4-141.8) | 255.7 (228.1-283.6) | 522.5 (450.2-595.2) |
| TB-associated mortality (per 100,000) | 25.9 (21.7-29.7) | 51.2 (44.5-58.9) | 106 (88.4-123) |
| TB notifications (per 100,000) | 54.6 (48.6-61.2) | 107.1 (94.2-120.5) | 221.3 (184.5-258.5) |
| Proportion aTB among infTB | 0.68 (0.65-0.71) | 0.68 (0.65-0.72) | 0.67 (0.64-0.71) |
| nTB:infTB ratio | 4.5 (3.9-5.1) | 4.5 (3.8-5.5) | 4.6 (3.6-5.5) |

### III. Screening implementation

Screening implementation required extending the base model (see Supplemental Materials Model Equations) to include four additional "on treatment via screening" compartments (see Figure 1), as well as a second parallel set of compartments and equations described in greater detail below.

For screening implementation, equations for the total population ($N(t)$), force of infection ($\lambda(t)$), and model entry ($\omega(t)$) were expanded to include relevant states from both sets of compartments.

$$N(t) = S(t) + I(t) + C(t) + Rn(t) + nTB(t) + aTB(t) + sTB(t) + Tp(t) + Ti(t) + Tc(t) + Tr(t) + Td(t) + Rt(t) + Sx(t) + Ix(t) + Cx(t) + Rnx(t) + nTBx(t) + aTBx(t) + sTBx(t) + Tpx(t) + Tix(t) + Tcx(t) + Trx(t) + Tdx(t) + Rtx(t)$$

$$\lambda(t) = \left(\beta * \frac{\left(t * aTB(t) + sTB(t) + t * aTBx(t) + sTBx(t)\right)}{N(t)}\right)$$

$$\omega(t) = \mu * N(t) + \mu_{sTB} * \left(sTB(t) + sTBx(t)\right)$$

The following set of equations describes the extended base model (here after referred to as "base compartments"). All individuals enter the model in this set of equations. During screening, these equations track individuals not yet considered for screening.

$$\frac{dS(t)}{dt} = -(\lambda(t) + \mu) * S(t) + \omega(t) + trtrec * Ti(t)$$

$$\frac{dC(t)}{dt} = -(\lambda(t) + \mu) * C(t) + infclr * I(t) + trtrec * Tc(t)$$

$$\frac{dRn(t)}{dt} = -(\lambda(t) * p + \mu) * Rn(t) + ntbrec * nTB(t) + trtrec * Tr(t)$$

$$\frac{dI(t)}{dt} = -(infclr + infntb + infatb + \mu) * I(t) + \lambda(t) * \left(S(t) + p * Rn(t) + r * Rt(t)\right)$$

$$\frac{dnTB(t)}{dt} = -(ntbrec + ntbatb + \mu) * nTB(t) + infntb * I(t) + atbntb * aTB(t)$$

$$\frac{daTB(t)}{dt} = -(atbntb + atbstb + \mu) * aTB(t) + infatb * I(t) + ntbatb * nTB(t) + stbatb * sTB(t)$$

$$\frac{dsTB(t)}{dt} = -(stbatb + stbtrt + \mu + \mu_{sTB}) * sTB(t) + atbstb * aTB(t)$$

$$\frac{dTp(t)}{dt} = -(trtrec + \mu) * Tp(t) + stbtrt * sTB(t)$$

$$\frac{dTi(t)}{dt} = -(trtrec + \mu) * Ti(t)$$

$$\frac{dTc(t)}{dt} = -(trtrec + \mu) * Tc(t)$$

$$\frac{dTr(t)}{dt} = -(trtrec + \mu) * Tr(t)$$

$$\frac{dTd(t)}{dt} = -(trtrec + \mu) * Td(t)$$

$$\frac{dRt(t)}{dt} = -(\lambda(t) * r + \mu) * Rt(t) + trtrec * Tp(t)$$

The following set of equations describes individuals who have been considered for screening (hereafter referred to as "screening compartments"). Compartments and equations are identical to the extended base model above, with "x" added to compartment names to distinguish the two sets of compartments and equations. During screening, these equations track individuals considered for screening.

$$\frac{dSx(t)}{dt} = -\lambda(t) * Sx(t) + trtrec * Tix(t)$$

$$\frac{dCx(t)}{dt} = -(\lambda(t) + \mu) * Cx(t) + infclr * Ix(t) + trtrec * Tcx(t)$$

$$\frac{dRnx(t)}{dt} = -(\lambda(t) * p + \mu) * Rnx(t) + ntbrec * nTBx(t) + trtrec * Trx(t)$$

$$\frac{dIx(t)}{dt} = -(infclr + infntb + infatb + \mu) * Ix(t) + \lambda(t) * \big(Sx(t) + p * Rnx(t) + r * Rtx(t)\big)$$

$$\frac{dnTBx(t)}{dt} = -(ntbrec + ntbatb + \mu) * nTBx(t) + infntb * Ix(t) + atbntb * aTBx(t)$$

$$\frac{daTBx(t)}{dt} = -(atbntb + atbstb + \mu) * aTBx(t) + infatb * Ix(t) + ntbatb * nTBx(t) + stbatb * sTBx(t)$$

$$\frac{dsTBx(t)}{dt} = -(stbatb + stbtrt + \mu + \mu_{sTB}) * sTBx(t) + atbstb * aTBx(t)$$

$$\frac{dTpx(t)}{dt} = -(trtrec + \mu) * Tpx(t) + stbtrt * sTBx(t)$$

$$\frac{dTix(t)}{dt} = -(trtrec + \mu) * Tix(t)$$

$$\frac{dTcx(t)}{dt} = -(trtrec + \mu) * Tcx(t)$$

$$\frac{dTrx(t)}{dt} = -(trtrec + \mu) * Trx(t)$$

$$\frac{dTdx(t)}{dt} = -(trtrec + \mu) * Tdx(t)$$

$$\frac{dRtx(t)}{dt} = -(\lambda(t) * r + \mu) * Rtx(t) + trtrec * Tpx(t)$$

Screening was implemented using the deSolve event function [8], which interrupts integration routines to allow changes in model states at designated timepoints. During each month of a 12-month round of screening, one-twelfth of each base compartment was considered eligible for screening and moved from base compartments to screening compartments using this function.

A proportion of these moved to one of four “on treatment via screening” compartments (Ts for Susceptible; Ti for Infection and Cleared; Tr for Recovered; Td for TB states and Previously treated), determined by population coverage and diagnostic algorithm. The diagnostic algorithm was defined using probability of a positive test for each tool. Probabilities of a positive test were broadly based on sensitivity for sTB and specificity for non-TB states [9-12], complemented by data from national TB prevalence surveys [13, 14], with assumptions where no data were available (see Supplementary Table 3 and Supplementary Table 4).

$$scr_x = population\ coverage * probabilty\ of\ a\ positive\ test_x$$

$$where\ x\ indicates\ the\ base\ compartment\ from\ which\ individuals\ were\ screened$$

The remaining proportion moved from base compartment to screening compartments without changing state (e.g., S to Sx). This included individuals not included in population coverage and those who were screened but tested negative.

At the end of a round of screening, all individuals in screening compartments were moved to base compartments, without changing state (e.g., Sx to S).

Supplemental Table 3: Summary of probability of a positive test result for each tool used in diagnostic algorithms by model state

| Model state | Probability of a positive test result | | | |
|---|---|---|---|---|
| | Prolonged cough | Chest x-ray | Xpert Ultra (screening use) | Xpert Ultra (confirmatory use) |
| Susceptible | 0.047-0.074 | 0.069-0.134 | 0.005-0.008 | 0.026-0.070 |
| Infected | 0.047-0.074 | 0.069-0.134 | 0.005-0.008 | 0.026-0.070 |
| Cleared | 0.047-0.074 | 0.069-0.134 | 0.005-0.008 | 0.026-0.070 |
| Recovered | 0.089-0.162 | 0.481-0.524 | 0.020-0.060 | 0.034-0.295 |
| Non-infectious TB | 0.129-0.249 | 0.626-0.712 | 0.026-0.070 | 0.026-0.070 |
| Asymptomatic infectious TB | 0.000 | 0.626-0.712 | 0.676-0.856 | 0.676-0.856 |
| Symptomatic infectious TB | 0.900-1.000 | 0.770-0.900 | 0.862-0.947 | 0.862-0.947 |
| Treated | 0.089-0.162 | 0.481-0.524 | 0.020-0.060 | 0.034-0.295 |

Supplemental Table 4: Probability of a positive test result for each screening diagnostic tool for each state in the model:

| Model state | Probability of a positive test range | Description | Reference |
|---|---|---|---|
| Prolonged cough | | | |
| Susceptible | 0.047-0.074 | Interquartile range for the proportion of national TB prevalence survey participants reporting prolonged (≥ 2 weeks) cough, regardless of CXR or TB status | [9, 13] |

| | | | |
|---|---|---|---|
| Infected | 0.047-0.074 | Interquartile range for the proportion of national TB prevalence survey participants reporting prolonged (≥ 2 weeks) cough, regardless of CXR or TB status | [9, 13] |
| Cleared | 0.047-0.074 | Interquartile range for the proportion of national TB prevalence survey participants reporting prolonged (≥ 2 weeks) cough, regardless of CXR or TB status | [9, 13] |
| Recovered | 0.089-0.162 | Midpoint between bounds for Susceptible/ Infected/Cleared and Non-infectious TB | Assumption |
| Non-infectious TB | 0.129-0.249 | Interquartile range for the proportion of national TB prevalence survey participants reporting prolonged (≥ 2 weeks) cough with abnormal CXR, regardless of TB status | [13] |
| Asymptomatic infectious TB | 0.00 | Assumption based on state definition | Assumption |
| Symptomatic infectious TB | 0.900-1.000 | Assumption based on state definition, with uncertainty recognising that individuals may report a symptom other than prolonged cough | Assumption |
| Treated | 0.089-0.162 | Midpoint between bounds for Susceptible/ Infected/Cleared and Non-infectious TB | Assumption |
| Chest X-ray | | | |
| Susceptible | 0.069-0.134 | Interquartile range for the proportion of national TB prevalence survey participants with abnormal CXR, regardless of TB status | [13] |
| Infected | 0.069-0.134 | Interquartile range for the proportion of national TB prevalence survey participants with abnormal CXR, regardless of TB status | [13] |
| Cleared | 0.069-0.134 | Interquartile range for the proportion of national TB prevalence survey participants with abnormal CXR, regardless of TB status | [13] |
| Recovered | 0.481-0.524 | Interquartile range for proportion with abnormal CXR suggestive of TB among participants of national TB prevalence survey reporting TB history | [14] |
| Non-infectious TB | 0.626-0.712 | Midpoint between the bounds for Recovered/Treated and Symptomatic infectious TB | Assumption |
| Asymptomatic infectious TB | 0.626-0.712 | Midpoint between the bounds for Recovered/Treated and Symptomatic infectious TB | Assumption |
| Symptomatic infectious TB | 0.770-0.900 | Sensitivity of CXR suggestive for TB for bacteriologically confirmed TB in screening use case | [9] |
| Treated | 0.481-0.524 | Interquartile range for proportion with abnormal CXR suggestive of TB among participants of national TB prevalence survey reporting TB history | [14] |
| Xpert Ultra: Screening use | | | |
| Susceptible | 0.005-0.008 | (1 – specificity) for individuals in a community in Kampala, Uganda | [11] |
| Infected | 0.005-0.008 | (1 – specificity) for individuals in a community in Kampala, Uganda | [11] |
| Cleared | 0.005-0.008 | (1 – specificity) for individuals in a community in Kampala, Uganda | [11] |

| | | | |
|---|---|---|---|
| Recovered | 0.020-0.060 | (1 – specificity) for individuals screened positive for symptoms and/or CXR with a history of TB in the community | [10] |
| Non-infectious TB | 0.026-0.070 | (1 – specificity) for pulmonary TB from individuals in primary care facilities and local hospitals | [12] |
| Asymptomatic infectious TB | 0.676-0.856 | Sensitivity for smear-negative TB from individuals in primary care facilities and local hospitals | [12] |
| Symptomatic infectious TB | 0.862-0.947 | Sensitivity for pulmonary TB from individuals in primary care facilities and local hospitals | [12] |
| Treated | 0.020-0.060 | (1 – specificity) for individuals screened positive for symptoms and/or CXR with a history of TB in the community | [10] |
| Xpert Ultra: Confirmatory use | | | |
| Susceptible | 0.026-0.070 | (1 – specificity) for pulmonary TB from individuals in primary care facilities and local hospitals | [12] |
| Infected | 0.026-0.070 | (1 – specificity) for pulmonary TB from individuals in primary care facilities and local hospitals | [12] |
| Cleared | 0.026-0.070 | (1 – specificity) for pulmonary TB from individuals in primary care facilities and local hospitals | [12] |
| Recovered | 0.034-0.295 | (1 – specificity) in individuals with history of TB treatment in primary care facilities and local hospitals | [12] |
| Non-infectious TB | 0.026-0.070 | (1 – specificity) for pulmonary TB from individuals in primary care facilities and local hospitals | [12] |
| Asymptomatic infectious TB | 0.676-0.856 | Sensitivity for smear-negative TB from individuals in primary care facilities and local hospitals | [12] |
| Symptomatic infectious TB | 0.862-0.947 | Sensitivity for pulmonary TB from individuals in primary care facilities and local hospitals | [12] |
| Treated | 0.034-0.295 | (1 – specificity) in individuals with history of TB treatment in primary care facilities and local hospitals | [12] |

### III. Results

Supplemental Table 5: Number of individuals who tested positive for TB in one round of screening with 100% coverage in a population with baseline prevalence of 500 per 100,000. sTB: symptomatic infectious TB; aTB: asymptomatic infectious TB; nTB: non-infectious TB; infectious TB: sTB+aTB; all TB: sTB+aTB+nTB.

| | Number of individuals with positive results | | |
|---|---|---|---|
| State | Cough+Xpert median (95% UI) | Xpert median (95% UI) | CXR median (95% UI) |
| sTB | 135 (115-158) | 142 (122-165) | 132 (112-153) |
| aTB | 0 (0-0) | 259 (222-298) | 226 (201-252) |
| nTB | 20 (10-36) | 66 (36-102) | 1,518 (1,220-1,875) |
| infectious TB | 135 (115-158) | 401 (359-446) | 359 (328-389) |
| all TB | 156 (132-184) | 469 (411-530) | 1,877 (1,567-2,242) |
| not TB | 443 (247-702) | 555 (418-717) | 13,516 (10,713-16,432) |

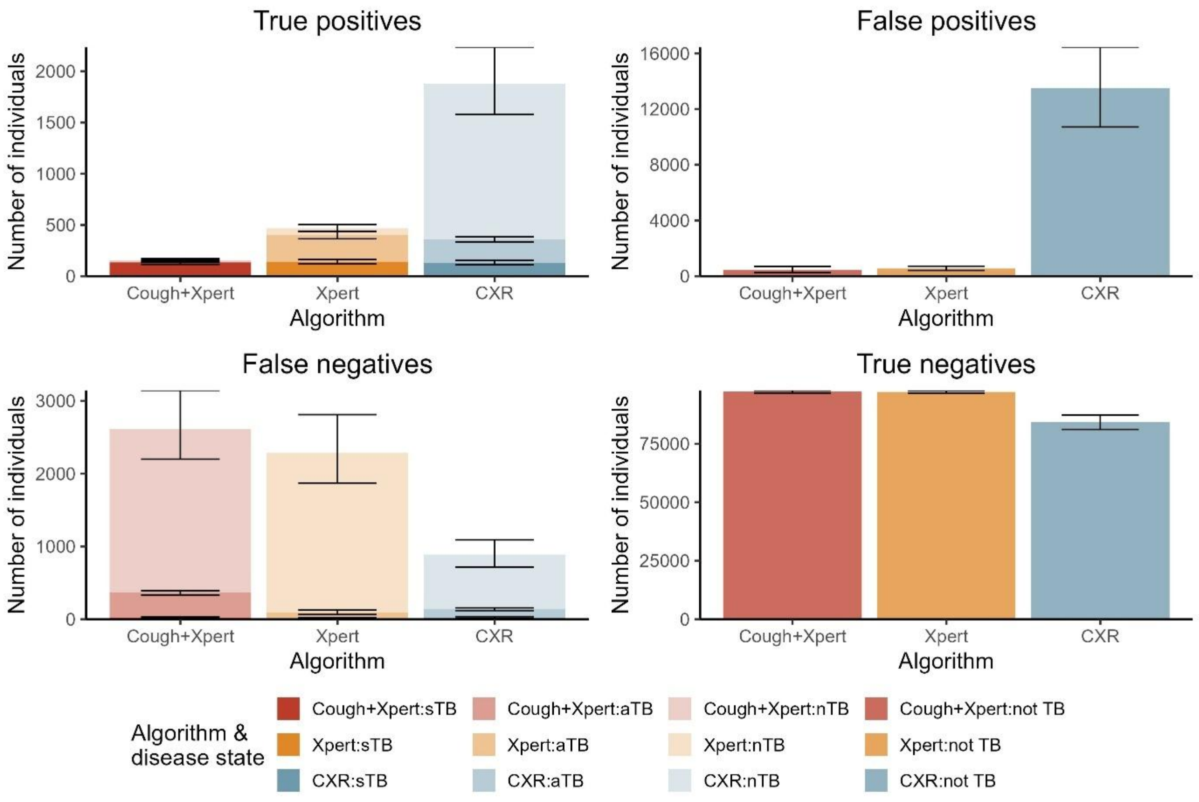


Supplemental Figure 1: Matrix showing number of individuals with true positive, false positive, false negative, and true negative results by diagnostic algorithm for one round of screening with 100% coverage in a population with baseline prevalence of 500 per 100,000. TB states are disaggregated to show symptomatic infectious TB (sTB), asymptomatic infectious TB (aTB), and non-infectious TB (nTB). Note y-axis differs in each quadrant.

Supplemental Table 6: Number of individuals with true positive results in each round of screening by diagnostic algorithm and population coverage for a population with baseline prevalence of 500 per 100,000.

| | Number of individuals with true positive results | | | | | | | | | |
|---|---|---|---|---|---|---|---|---|---|---|
| | Population coverage | | | | | | | | | |
| Round | 10% | 20% | 30% | 40% | 50% | 60% | 70% | 80% | 90% | 100% |
| Algorithm targeting symptomatic infectious TB (Cough+Xpert) | | | | | | | | | | |
| 1 | 16 (13-19) | 31 (27-37) | 47 (40-55) | 63 (53-74) | 78 (66-93) | 93 (79-111) | 109 (91-128) | 125 (106-150) | 141 (118-169) | 156 (133-187) |
| 2 | 15 (13-18) | 30 (25-35) | 44 (37-51) | 57 (48-67) | 69 (58-82) | 81 (69-95) | 92 (78-107) | 102 (88-122) | 112 (95-133) | 121 (103-142) |
| 3 | 15 (13-18) | 29 (25-34) | 42 (36-50) | 55 (46-64) | 66 (56-78) | 77 (65-90) | 87 (74-101) | 96 (83-114) | 105 (89-124) | 112 (96-132) |
| 4 | 15 (13-18) | 29 (25-34) | 41 (36-49) | 53 (45-63) | 64 (54-75) | 74 (63-86) | 83 (71-96) | 91 (79-108) | 99 (85-117) | 106 (91-124) |
| 5 | 15 (13-18) | 29 (24-33) | 41 (35-48) | 52 (44-61) | 62 (53-73) | 71 (61-83) | 79 (68-92) | 87 (75-103) | 94 (81-110) | 100 (87-116) |
| Algorithm targeting infectious TB (Xpert) | | | | | | | | | | |
| 1 | 47 (41-54) | 95 (82-107) | 142 (124-160) | 188 (165-213) | 236 (205-267) | 282 (247-318) | 326 (290-375) | 375 (331-426) | 421 (365-475) | 468 (405-527) |
| 2 | 46 (40-52) | 88 (77-100) | 128 (112-144) | 163 (143-185) | 196 (170-223) | 225 (198-255) | 250 (220-288) | 274 (240-315) | 294 (255-339) | 311 (269-357) |
| 3 | 44 (39-51) | 84 (73-95) | 119 (104-134) | 148 (129-168) | 174 (152-200) | 195 (172-223) | 213 (186-245) | 228 (199-264) | 241 (208-282) | 250 (215-293) |
| 4 | 43 (38-50) | 80 (70-91) | 111 (97-126) | 136 (119-155) | 156 (136-180) | 172 (151-198) | 185 (161-214) | 194 (167-226) | 201 (171-238) | 206 (175-243) |
| 5 | 43 (38-49) | 77 (67-88) | 104 (92-118) | 125 (109-143) | 141 (122-164) | 152 (133-177) | 161 (139-187) | 165 (141-195) | 168 (142-202) | 168 (142-203) |
| Algorithm targeting all TB (CXR) | | | | | | | | | | |
| 1 | 188 (156-226) | 378 (312-453) | 567 (467-680) | 753 (619-914) | 937 (782-1132) | 1124 (933-1367) | 1309 (1099-1601) | 1497 (1233-1820) | 1688 (1410-2032) | 1872 (1556-2255) |
| 2 | 176 (146-212) | 329 (274-395) | 458 (380-553) | 562 (465-681) | 640 (536-768) | 696 (586-837) | 728 (611-879) | 738 (617-885) | 723 (608-869) | 687 (574-828) |
| 3 | 165 (137-198) | 289 (240-346) | 371 (309-448) | 421 (351-508) | 441 (368-530) | 436 (366-523) | 411 (343-494) | 370 (309-448) | 319 (261-394) | 262 (212-327) |
| 4 | 155 (129-186) | 253 (211-304) | 303 (253-364) | 317 (263-381) | 305 (256-369) | 275 (230-329) | 234 (192-284) | 188 (153-233) | 142 (113-182) | 101 (77-133) |
| 5 | 146 (121-175) | 222 (185-268) | 248 (207-298) | 240 (200-289) | 212 (177-257) | 175 (144-211) | 133 (108-166) | 96 (76-122) | 63 (48-86) | 39 (28-55) |

Supplemental Table 7: Number of individuals with false positive results in each round of screening by diagnostic algorithm and population coverage for a population with baseline prevalence of 500 per 100,000.

| | Number of individuals with false positive results | | | | | | | | | |
|---|---|---|---|---|---|---|---|---|---|---|
| | Population coverage | | | | | | | | | |
| Round | 10% | 20% | 30% | 40% | 50% | 60% | 70% | 80% | 90% | 100% |
| Algorithm targeting symptomatic infectious TB (Cough+Xpert) | | | | | | | | | | |
| 1 | 44<br>(23-67) | 87<br>(48-135) | 132<br>(70-202) | 174<br>(93-277) | 225<br>(116-349) | 265<br>(138-414) | 306<br>(168-468) | 353<br>(189-559) | 391<br>(221-611) | 436<br>(235-685) |
| 2 | 44<br>(23-67) | 87<br>(48-135) | 132<br>(70-202) | 174<br>(93-277) | 226<br>(116-350) | 266<br>(139-415) | 307<br>(169-469) | 354<br>(190-560) | 392<br>(222-613) | 439<br>(237-688) |
| 3 | 44<br>(23-67) | 87<br>(48-135) | 132<br>(70-202) | 175<br>(93-278) | 226<br>(116-351) | 266<br>(139-416) | 308<br>(169-470) | 356<br>(191-562) | 394<br>(223-616) | 441<br>(239-690) |
| 4 | 44<br>(23-67) | 87<br>(48-135) | 132<br>(70-202) | 175<br>(93-278) | 227<br>(116-352) | 267<br>(139-417) | 308<br>(170-471) | 357<br>(191-563) | 395<br>(224-619) | 443<br>(240-692) |
| 5 | 44<br>(23-67) | 87<br>(48-135) | 132<br>(70-203) | 175<br>(93-278) | 227<br>(116-353) | 268<br>(140-418) | 309<br>(170-471) | 358<br>(191-564) | 396<br>(224-620) | 444<br>(241-694) |
| Algorithm targeting infectious TB (Xpert) | | | | | | | | | | |
| 1 | 57<br>(42-72) | 112<br>(85-143) | 169<br>(125-214) | 223<br>(165-289) | 280<br>(212-364) | 334<br>(249-441) | 393<br>(300-503) | 448<br>(342-570) | 500<br>(378-653) | 559<br>(421-716) |
| 2 | 57<br>(42-72) | 112<br>(85-144) | 169<br>(126-215) | 224<br>(167-290) | 281<br>(214-367) | 337<br>(252-445) | 398<br>(302-509) | 454<br>(346-577) | 509<br>(385-661) | 569<br>(428-725) |
| 3 | 57<br>(42-72) | 113<br>(85-144) | 170<br>(126-216) | 226<br>(168-291) | 284<br>(216-369) | 340<br>(254-448) | 401<br>(305-513) | 459<br>(348-583) | 515<br>(391-669) | 575<br>(436-732) |
| 4 | 57<br>(42-73) | 113<br>(86-144) | 171<br>(127-217) | 227<br>(169-292) | 285<br>(217-370) | 342<br>(255-450) | 404<br>(308-516) | 462<br>(350-587) | 519<br>(394-674) | 580<br>(439-738) |
| 5 | 57<br>(42-73) | 113<br>(86-145) | 171<br>(127-217) | 227<br>(169-293) | 286<br>(218-372) | 343<br>(256-452) | 405<br>(309-517) | 464<br>(351-590) | 522<br>(396-677) | 583<br>(441-743) |
| Algorithm targeting all TB (CXR) | | | | | | | | | | |
| 1 | 1372<br>(1070-1645) | 2714<br>(2135-3269) | 4053<br>(3223-4912) | 5436<br>(4313-6518) | 6842<br>(5406-8208) | 8049<br>(6431-9822) | 9497<br>(7496-11457) | 10911<br>(8536-13057) | 12316<br>(9734-14777) | 13592<br>(10620-16510) |
| 2 | 1372<br>(1071-1646) | 2716<br>(2142-3270) | 4056<br>(3221-4911) | 5437<br>(4320-6515) | 6859<br>(5421-8208) | 8090<br>(6473-9841) | 9516<br>(7555-11478) | 10958<br>(8621-13078) | 12393<br>(9832-14816) | 13676<br>(10766-16555) |
| 3 | 1376<br>(1074-1651) | 2735<br>(2155-3290) | 4091<br>(3256-4953) | 5502<br>(4369-6586) | 6946<br>(5499-8301) | 8205<br>(6576-9993) | 9668<br>(7702-11664) | 11135<br>(8768-13308) | 12613<br>(10022-15067) | 13899<br>(10943-16816) |
| 4 | 1379<br>(1077-1655) | 2746<br>(2165-3306) | 4117<br>(3275-4985) | 5547<br>(4393-6635) | 7004<br>(5545-8363) | 8276<br>(6643-10085) | 9739<br>(7771-11762) | 11222<br>(8847-13428) | 12695<br>(10077-15172) | 13975<br>(10992-16910) |
| 5 | 1380<br>(1078-1658) | 2754<br>(2173-3319) | 4132<br>(3280-5005) | 5571<br>(4405-6665) | 7031<br>(5564-8396) | 8307<br>(6677-10123) | 9771<br>(7796-11805) | 11247<br>(8859-13457) | 12702<br>(10075-15176) | 13976<br>(10979-16903) |

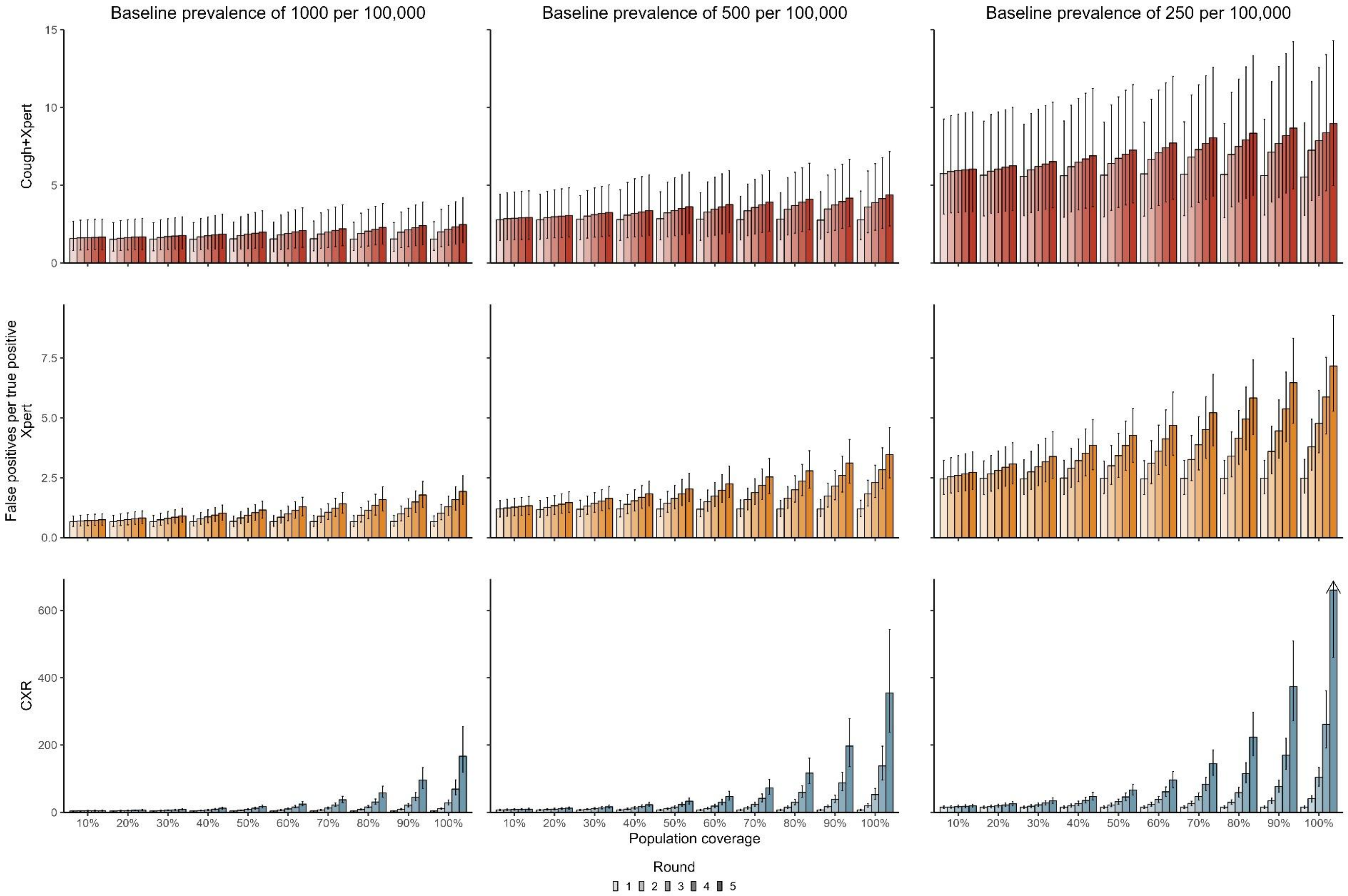


Supplemental Figure 2: Number of individuals with false positive results per individual with true positive results for each round of screening by diagnostic algorithm, population coverage, and baseline prevalence.

Supplemental Table 8: Number of individuals with false positive results per individual with true positive results for each round of screening by diagnostic algorithm and population coverage for a population with baseline prevalence of 500 per 100,000.

| | Number of individuals with false positive results per individual with true positive results | | | | | | | | | |
|---|---|---|---|---|---|---|---|---|---|---|
| | Population coverage | | | | | | | | | |
| Round | 10% | 20% | 30% | 40% | 50% | 60% | 70% | 80% | 90% | 100% |
| Algorithm targeting symptomatic infectious TB (Cough+Xpert) | | | | | | | | | | |
| 1 | 2.8 (1.4-4.4) | 2.8 (1.5-4.4) | 2.8 (1.5-4.3) | 2.8 (1.5-4.7) | 2.8 (1.5-4.6) | 2.8 (1.5-4.5) | 2.8 (1.5-4.3) | 2.8 (1.5-4.5) | 2.8 (1.5-4.6) | 2.8 (1.5-4.6) |
| 2 | 2.9 (1.5-4.5) | 2.9 (1.6-4.6) | 3.0 (1.6-4.7) | 3.1 (1.6-5.2) | 3.2 (1.7-5.2) | 3.3 (1.7-5.2) | 3.4 (1.8-5.1) | 3.5 (1.8-5.5) | 3.5 (2.0-5.6) | 3.6 (1.9-5.9) |
| 3 | 2.9 (1.5-4.6) | 3.0 (1.6-4.7) | 3.1 (1.7-4.8) | 3.2 (1.7-5.4) | 3.4 (1.8-5.5) | 3.5 (1.8-5.5) | 3.6 (1.9-5.4) | 3.7 (1.9-5.8) | 3.7 (2.1-6.0) | 3.9 (2.1-6.4) |
| 4 | 2.9 (1.5-4.6) | 3.0 (1.6-4.8) | 3.2 (1.7-4.9) | 3.3 (1.7-5.6) | 3.5 (1.8-5.7) | 3.6 (1.9-5.7) | 3.7 (2.0-5.7) | 3.9 (2.0-6.1) | 4.0 (2.2-6.3) | 4.1 (2.2-6.8) |
| 5 | 2.9 (1.5-4.6) | 3.1 (1.7-4.8) | 3.2 (1.7-5.0) | 3.4 (1.8-5.7) | 3.6 (1.9-5.8) | 3.8 (2.0-5.9) | 3.9 (2.1-5.9) | 4.1 (2.1-6.4) | 4.2 (2.4-6.7) | 4.4 (2.4-7.2) |
| Algorithm targeting infectious TB (Xpert) | | | | | | | | | | |
| 1 | 1.2 (0.9-1.5) | 1.2 (0.9-1.6) | 1.2 (0.9-1.6) | 1.2 (0.9-1.6) | 1.2 (0.9-1.6) | 1.2 (0.9-1.6) | 1.2 (0.9-1.6) | 1.2 (0.9-1.6) | 1.2 (0.9-1.6) | 1.2 (0.9-1.6) |
| 2 | 1.2 (0.9-1.6) | 1.3 (0.9-1.7) | 1.3 (1.0-1.7) | 1.4 (1.0-1.8) | 1.4 (1.1-1.9) | 1.5 (1.1-2.0) | 1.6 (1.2-2.1) | 1.7 (1.2-2.1) | 1.7 (1.3-2.3) | 1.8 (1.3-2.4) |
| 3 | 1.3 (0.9-1.6) | 1.3 (1.0-1.8) | 1.4 (1.1-1.9) | 1.5 (1.1-2.0) | 1.6 (1.2-2.2) | 1.7 (1.3-2.3) | 1.9 (1.4-2.5) | 2.0 (1.5-2.6) | 2.2 (1.6-2.8) | 2.3 (1.7-3.0) |
| 4 | 1.3 (0.9-1.7) | 1.4 (1.0-1.8) | 1.5 (1.1-2.0) | 1.7 (1.2-2.2) | 1.8 (1.3-2.4) | 2.0 (1.5-2.6) | 2.2 (1.6-2.9) | 2.4 (1.8-3.1) | 2.6 (1.9-3.4) | 2.8 (2.0-3.8) |
| 5 | 1.3 (1.0-1.7) | 1.5 (1.1-1.9) | 1.6 (1.2-2.1) | 1.8 (1.3-2.4) | 2.0 (1.5-2.7) | 2.3 (1.7-3.0) | 2.5 (1.8-3.3) | 2.8 (2.1-3.6) | 3.1 (2.3-4.1) | 3.5 (2.5-4.6) |
| Algorithm targeting all TB (CXR) | | | | | | | | | | |
| 1 | 7.2 (5.4-9.4) | 7.2 (5.4-9.3) | 7.1 (5.4-9.3) | 7.1 (5.4-9.4) | 7.3 (5.4-9.3) | 7.1 (5.4-9.4) | 7.2 (5.4-9.4) | 7.2 (5.4-9.4) | 7.2 (5.5-9.3) | 7.2 (5.5-9.4) |
| 2 | 7.7 (5.7-10.1) | 8.2 (6.3-10.6) | 8.8 (6.7-11.4) | 9.6 (7.4-12.5) | 10.7 (8.0-13.5) | 11.5 (8.8-15.1) | 13.0 (9.9-16.8) | 14.7 (11.2-19.0) | 16.9 (12.9-21.8) | 19.7 (15.2-25.5) |
| 3 | 8.3 (6.1-10.7) | 9.5 (7.2-12.2) | 10.9 (8.3-14.1) | 12.8 (9.9-16.8) | 15.8 (11.7-20.0) | 18.7 (14.3-24.3) | 23.4 (17.9-30.5) | 29.7 (22.7-38.9) | 38.9 (29.5-50.9) | 52.7 (38.8-71.1) |
| 4 | 8.8 (6.6-11.4) | 10.8 (8.3-14.0) | 13.5 (10.3-17.4) | 17.1 (13.3-22.5) | 23 (17.3-29.0) | 29.9 (22.9-39.1) | 41.6 (31.3-54.7) | 59.2 (44.4-78.7) | 88.1 (63.9-119.4) | 137.7 (96.7-196.7) |
| 5 | 9.4 (7.0-12.2) | 12.3 (9.4-15.9) | 16.5 (12.6-21.3) | 22.7 (17.6-30.0) | 33.1 (24.8-42.3) | 47.3 (35.9-62.7) | 72.9 (53.6-97.7) | 116.5 (84.9-160.8) | 196.9 (135.6-278.5) | 354.9 (237.7-543.3) |

Supplemental Table 9: Number of individuals with false positive results per individual with true positive results for each round of screening by diagnostic algorithm and population coverage for a population with baseline prevalence of 1,000 per 100,000.

| | Number of individuals with false positive results per individual with true positive results | | | | | | | | | |
|---|---|---|---|---|---|---|---|---|---|---|
| | Population coverage | | | | | | | | | |
| Round | 10% | 20% | 30% | 40% | 50% | 60% | 70% | 80% | 90% | 100% |
| Algorithm targeting symptomatic infectious TB (Cough+Xpert) | | | | | | | | | | |
| 1 | 1.6 (0.8-2.7) | 1.5 (0.8-2.6) | 1.5 (0.8-2.6) | 1.5 (0.8-2.6) | 1.6 (0.8-2.6) | 1.6 (0.7-2.6) | 1.6 (0.8-2.7) | 1.5 (0.8-2.6) | 1.5 (0.8-2.6) | 1.5 (0.8-2.7) |
| 2 | 1.6 (0.8-2.8) | 1.6 (0.8-2.7) | 1.6 (0.8-2.8) | 1.7 (0.8-2.9) | 1.8 (0.9-3.0) | 1.8 (0.9-3.1) | 1.9 (0.9-3.2) | 1.9 (1.0-3.2) | 2.0 (1.0-3.3) | 2.0 (1.1-3.4) |
| 3 | 1.6 (0.8-2.8) | 1.6 (0.8-2.8) | 1.7 (0.9-2.8) | 1.8 (0.9-3.0) | 1.9 (0.9-3.1) | 1.9 (0.9-3.3) | 2.0 (1.0-3.4) | 2.1 (1.1-3.5) | 2.1 (1.1-3.5) | 2.2 (1.1-3.7) |
| 4 | 1.6 (0.8-2.8) | 1.7 (0.8-2.8) | 1.7 (0.9-2.9) | 1.8 (0.9-3.1) | 1.9 (1.0-3.2) | 2.0 (1.0-3.4) | 2.1 (1.1-3.6) | 2.2 (1.2-3.6) | 2.3 (1.1-3.7) | 2.3 (1.2-3.9) |
| 5 | 1.7 (0.9-2.8) | 1.7 (0.9-2.9) | 1.8 (0.9-3.0) | 1.9 (0.9-3.1) | 2.0 (1.0-3.4) | 2.1 (1.0-3.5) | 2.2 (1.1-3.7) | 2.3 (1.2-3.8) | 2.4 (1.2-3.9) | 2.5 (1.3-4.2) |
| Algorithm targeting infectious TB (Xpert) | | | | | | | | | | |
| 1 | 0.7 (0.5-0.9) | 0.7 (0.5-0.9) | 0.7 (0.5-0.9) | 0.7 (0.5-0.9) | 0.7 (0.5-0.9) | 0.7 (0.5-0.9) | 0.7 (0.5-0.9) | 0.7 (0.5-0.9) | 0.7 (0.5-0.9) | 0.7 (0.5-0.9) |
| 2 | 0.7 (0.5-0.9) | 0.7 (0.5-1.0) | 0.7 (0.5-1.0) | 0.8 (0.6-1.1) | 0.8 (0.6-1.1) | 0.9 (0.6-1.1) | 0.9 (0.6-1.2) | 0.9 (0.7-1.3) | 1.0 (0.7-1.3) | 1.0 (0.7-1.4) |
| 3 | 0.7 (0.5-1.0) | 0.8 (0.5-1.0) | 0.8 (0.6-1.1) | 0.9 (0.6-1.2) | 0.9 (0.7-1.2) | 1 (0.7-1.3) | 1.1 (0.8-1.4) | 1.1 (0.8-1.5) | 1.2 (0.9-1.6) | 1.3 (0.9-1.7) |
| 4 | 0.7 (0.5-1.0) | 0.8 (0.6-1.1) | 0.9 (0.6-1.2) | 0.9 (0.7-1.3) | 1.0 (0.7-1.4) | 1.1 (0.8-1.5) | 1.2 (0.9-1.6) | 1.4 (0.9-1.8) | 1.5 (1.1-2.0) | 1.6 (1.2-2.1) |
| 5 | 0.7 (0.5-1.0) | 0.8 (0.6-1.1) | 0.9 (0.7-1.2) | 1.0 (0.7-1.4) | 1.2 (0.8-1.5) | 1.3 (0.9-1.7) | 1.4 (1.0-1.9) | 1.6 (1.1-2.1) | 1.8 (1.3-2.4) | 1.9 (1.4-2.6) |
| Algorithm targeting all TB (CXR) | | | | | | | | | | |
| 1 | 3.9 (3.1-5.0) | 3.9 (3.1-5.0) | 4.0 (3.0-5.0) | 3.9 (3.1-5.0) | 3.9 (3.1-4.9) | 3.9 (3.1-5.0) | 4.0 (3.1-5.0) | 3.9 (3.1-5.0) | 3.9 (3.0-5.0) | 4.0 (3.1-5.0) |
| 2 | 4.2 (3.3-5.3) | 4.5 (3.5-5.8) | 4.9 (3.8-6.2) | 5.3 (4.2-6.7) | 5.8 (4.6-7.2) | 6.4 (5.1-8.1) | 7.2 (5.7-9.0) | 8.1 (6.5-10.1) | 9.3 (7.4-11.7) | 11.0 (8.8-13.8) |
| 3 | 4.5 (3.5-5.7) | 5.2 (4.0-6.6) | 6.1 (4.7-7.6) | 7.2 (5.6-8.9) | 8.5 (6.8-10.6) | 10.2 (8.3-12.9) | 12.8 (10.2-15.9) | 16.1 (13.0-20.4) | 21.0 (16.5-26.5) | 28.3 (22.4-37.1) |
| 4 | 4.8 (3.7-6.0) | 5.9 (4.6-7.5) | 7.4 (5.8-9.3) | 9.5 (7.4-11.7) | 12.2 (9.8-15.1) | 16 (13.0-20.4) | 22.1 (17.6-27.5) | 31.0 (24.5-39.7) | 45.2 (34.9-58.8) | 69.3 (52.4-96.7) |
| 5 | 5.1 (4.0-6.4) | 6.6 (5.2-8.4) | 8.9 (7.0-11.2) | 12.3 (9.7-15.2) | 17.3 (13.8-21.5) | 24.7 (19.9-31.6) | 37.5 (29.2-47.8) | 58.4 (44.8-77.7) | 96.1 (70.7-132.9) | 166.9 (118.8-254.4) |

Supplemental Table 10: Number of individuals with false positive results per individual with true positive results for each round of screening by diagnostic algorithm and population coverage for a population with infectious prevalence of 250 per 100,000.

| | Number of individuals with false positive results per individual with true positive results | | | | | | | | | |
|---|---|---|---|---|---|---|---|---|---|---|
| | Population coverage | | | | | | | | | |
| Round | 10% | 20% | 30% | 40% | 50% | 60% | 70% | 80% | 90% | 100% |
| Algorithm targeting symptomatic infectious TB (Cough+Xpert) | | | | | | | | | | |
| 1 | 5.7 (3.1-9.3) | 5.6 (3.0-9.1) | 5.6 (3.0-8.9) | 5.6 (2.9-9.1) | 5.6 (3.0-9.1) | 5.7 (3.0-9.0) | 5.7 (3.0-9.1) | 5.7 (2.9-9.0) | 5.6 (3.1-9.2) | 5.5 (3.1-9.0) |
| 2 | 5.9 (3.2-9.5) | 5.9 (3.2-9.5) | 6.0 (3.3-9.6) | 6.2 (3.3-10.1) | 6.4 (3.4-10.2) | 6.7 (3.5-10.5) | 6.8 (3.6-10.8) | 7.0 (3.6-11.0) | 7.1 (3.9-11.7) | 7.2 (4.0-11.7) |
| 3 | 5.9 (3.3-9.6) | 6.0 (3.3-9.7) | 6.2 (3.4-9.9) | 6.5 (3.4-10.6) | 6.7 (3.6-10.7) | 7.1 (3.7-11.1) | 7.3 (3.9-11.5) | 7.5 (3.9-11.8) | 7.7 (4.2-12.6) | 7.9 (4.3-12.6) |
| 4 | 6.0 (3.3-9.7) | 6.2 (3.3-9.9) | 6.4 (3.5-10.1) | 6.7 (3.5-10.9) | 7.0 (3.7-11.1) | 7.4 (3.9-11.6) | 7.7 (4.1-12.0) | 7.9 (4.1-12.6) | 8.2 (4.5-13.5) | 8.4 (4.6-13.4) |
| 5 | 6.0 (3.3-9.7) | 6.2 (3.4-10.0) | 6.5 (3.5-10.3) | 6.9 (3.6-11.2) | 7.3 (3.9-11.5) | 7.7 (4.1-12.0) | 8.1 (4.3-12.6) | 8.3 (4.4-13.3) | 8.7 (4.8-14.2) | 9.0 (5.0-14.3) |
| Algorithm targeting infectious TB (Xpert) | | | | | | | | | | |
| 1 | 2.5 (1.8-3.2) | 2.5 (1.8-3.2) | 2.5 (1.8-3.2) | 2.5 (1.8-3.2) | 2.5 (1.8-3.2) | 2.5 (1.8-3.2) | 2.5 (1.8-3.2) | 2.5 (1.8-3.2) | 2.5 (1.8-3.2) | 2.5 (1.9-3.3) |
| 2 | 2.5 (1.9-3.3) | 2.7 (1.9-3.4) | 2.7 (2.0-3.6) | 2.9 (2.1-3.7) | 3.0 (2.2-3.8) | 3.1 (2.3-4.1) | 3.3 (2.4-4.3) | 3.4 (2.5-4.4) | 3.6 (2.7-4.7) | 3.8 (2.8-4.9) |
| 3 | 2.6 (1.9-3.4) | 2.8 (2.0-3.6) | 3.0 (2.2-3.9) | 3.2 (2.3-4.1) | 3.4 (2.5-4.4) | 3.6 (2.6-4.7) | 3.9 (2.8-5.1) | 4.2 (3.1-5.3) | 4.5 (3.3-5.8) | 4.8 (3.5-6.2) |
| 4 | 2.7 (2.0-3.5) | 2.9 (2.1-3.8) | 3.2 (2.3-4.2) | 3.5 (2.6-4.5) | 3.8 (2.8-4.9) | 4.1 (3.0-5.3) | 4.5 (3.3-5.9) | 5.0 (3.7-6.3) | 5.4 (4.0-6.9) | 5.9 (4.3-7.5) |
| 5 | 2.7 (2.0-3.6) | 3.1 (2.2-4.0) | 3.4 (2.5-4.4) | 3.9 (2.8-4.9) | 4.3 (3.1-5.4) | 4.7 (3.4-6.1) | 5.2 (3.8-6.8) | 5.8 (4.3-7.4) | 6.5 (4.8-8.3) | 7.2 (5.3-9.3) |
| Algorithm targeting all TB (CXR) | | | | | | | | | | |
| 1 | 14.8 (11.6-18.8) | 14.8 (11.2-18.7) | 14.8 (11.6-18.7) | 14.9 (11.6-19.0) | 15.0 (11.5-18.9) | 14.9 (11.4-18.8) | 15.0 (11.5-18.9) | 14.8 (11.5-19.1) | 15.1 (11.7-18.8) | 15.3 (11.6-18.9) |
| 2 | 15.7 (12.4-19.9) | 16.9 (12.9-21.3) | 18.2 (14.3-22.8) | 19.8 (15.4-25.0) | 21.7 (16.8-27.3) | 23.8 (18.4-29.7) | 26.5 (20.3-33.1) | 29.4 (23.3-37.3) | 34.2 (26.7-42.1) | 40.3 (30.9-49.6) |
| 3 | 16.8 (13.3-21.3) | 19.4 (15.0-24.4) | 22.5 (17.7-28.2) | 26.4 (20.6-33.3) | 31.6 (24.5-39.4) | 38.2 (29.5-47.5) | 47.1 (36.3-58.3) | 58.6 (45.8-73.9) | 76.7 (59.4-95.7) | 104.1 (77.6-133.7) |
| 4 | 18.0 (14.2-22.7) | 22.2 (17.2-27.9) | 27.8 (21.9-34.9) | 35.2 (27.7-44.5) | 46.0 (35.5-57.2) | 61.1 (46.8-75.7) | 83.0 (63.8-103.6) | 114.6 (88.6-147.6) | 169.7 (127.8-219.9) | 261.9 (190.8-361.5) |
| 5 | 19.1 (15.1-24.2) | 25.4 (19.7-31.9) | 34.0 (26.9-42.9) | 46.8 (36.6-59.0) | 66.5 (51.3-83.0) | 96.6 (73.4-121.3) | 144.9 (110.0-185.7) | 223.8 (168.3-296.9) | 374.1 (271.3-509.0) | 660.6 (460.2-974.2) |

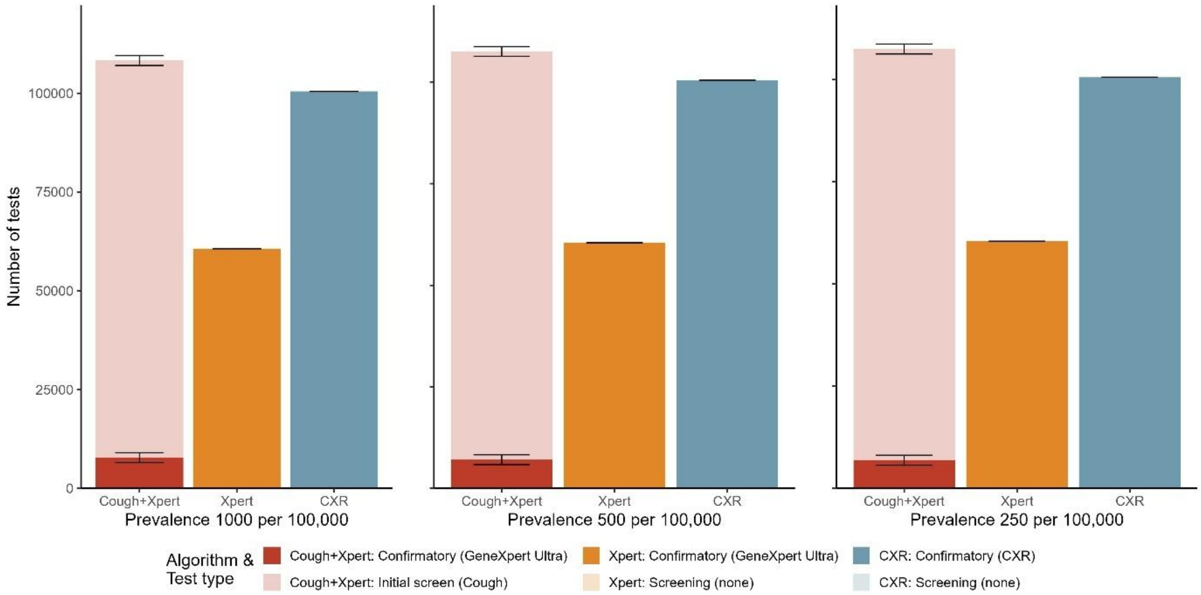


Supplemental Figure 3: Number of initial screen (light colour) and confirmatory (dark colour) tests required for one round of screening with 100% coverage by diagnostic algorithm in populations with different baseline prevalence levels.

Supplemental Table 11: Projected percentage of symptomatic infectious TB episodes averted (2025-2035) by diagnostic algorithm, population coverage, and duration for a population with baseline prevalence of 500 per 100,000

| | Projected reduction in symptomatic infectious TB episodes (2025-2035) | | | | | | | | | |
|---|---|---|---|---|---|---|---|---|---|---|
| | Population coverage | | | | | | | | | |
| Rounds | 10% | 20% | 30% | 40% | 50% | 60% | 70% | 80% | 90% | 100% |
| Algorithm targeting symptomatic infectious TB (Cough+Xpert) | | | | | | | | | | |
| 1 | 0.6% (0.5-0.7%) | 1.1% (0.9-1.4%) | 1.7% (1.4-2.0%) | 2.3% (1.9-2.7%) | 2.8% (2.3-3.4%) | 3.4% (2.8-4.1%) | 3.9% (3.2-4.8%) | 4.5% (3.7-5.5%) | 5.1% (4.2-6.1%) | 5.6% (4.7-6.8%) |
| 2 | 1.1% (0.9-1.3%) | 2.1% (1.7-2.5%) | 3.1% (2.6-3.7%) | 4.1% (3.4-5.0%) | 5.1% (4.2-6.1%) | 6.0% (5.0-7.2%) | 6.9% (5.8-8.3%) | 7.8% (6.6-9.3%) | 8.7% (7.3-10.5%) | 9.6% (7.9-11.5%) |
| 3 | 1.5% (1.2-1.8%) | 2.9% (2.4-3.5%) | 4.3% (3.6-5.2%) | 5.7% (4.7-6.9%) | 7.0% (5.8-8.3%) | 8.2% (6.9-9.8%) | 9.4% (7.9-11.2%) | 10.6% (8.9-12.8%) | 11.8% (9.9-13.9%) | 12.8% (10.8-15.3%) |
| 4 | 1.9% (1.5-2.3%) | 3.7% (3.1-4.4%) | 5.4% (4.5-6.5%) | 7.0% (5.8-8.4%) | 8.6% (7.2-10.3%) | 10.1% (8.5-12.0%) | 11.6% (9.7-13.8%) | 12.9% (10.9-15.4%) | 14.2% (12.1-16.9%) | 15.6% (13.0-18.4%) |
| 5 | 2.2% (1.8-2.7%) | 4.3% (3.5-5.2%) | 6.3% (5.2-7.5%) | 8.2% (6.8-9.7%) | 9.9% (8.3-11.8%) | 11.6% (9.9-13.9%) | 13.3% (11.3-15.7%) | 14.9% (12.4-17.8%) | 16.4% (13.9-19.3%) | 17.8% (15.1-20.9%) |
| Algorithm targeting infectious TB (Xpert) | | | | | | | | | | |
| 1 | 1.9% (1.7-2.3%) | 3.9% (3.3-4.5%) | 5.9% (5-6.8%) | 7.8% (6.6-8.9%) | 9.8% (8.4-11.3%) | 11.8% (10.1-13.6%) | 13.7% (11.9-15.9%) | 15.8% (13.4-18.2%) | 17.7% (15.1-20.6%) | 19.6% (16.9-22.8%) |
| 2 | 3.7% (3.1-4.2%) | 7.2% (6.3-8.3%) | 10.7% (9.2-12.4%) | 14% (12.2-16.2%) | 17.4% (15.1-19.9%) | 20.5% (17.8-23.4%) | 23.5% (20.4-26.9%) | 26.5% (22.9-30.0%) | 29.3% (25.5-33.3%) | 32% (28-36.1%) |
| 3 | 5.2% (4.5-6.0%) | 10.2% (8.7-11.7%) | 14.8% (12.8-17.0%) | 19.3% (16.7-21.9%) | 23.5% (20.5-26.7%) | 27.5% (23.9-30.9%) | 31.2% (27.5-34.9%) | 34.8% (30.4-39%) | 38.0% (33.2-42.6%) | 41.0% (36.3-45.8%) |
| 4 | 6.5% (5.6-7.5%) | 12.6% (11.0-14.4%) | 18.2% (15.8-20.9%) | 23.5% (20.6-26.7%) | 28.4% (24.8-32.1%) | 32.9% (29.0-37.1%) | 37.2% (33.1-41.4%) | 40.9% (36.0-45.5%) | 44.6% (39.9-49.2%) | 48.0% (42.9-52.5%) |
| 5 | 7.7% (6.6-8.8%) | 14.7% (12.8-16.7%) | 21.2% (18.3-24.1%) | 27.0% (23.6-30.3%) | 32.4% (28.5-36.3%) | 37.1% (33.1-41.2%) | 41.5% (36.9-46.4%) | 45.6% (40.9-50.2%) | 49.3% (44.3-53.9%) | 52.7% (47.9-57.5%) |
| Algorithm targeting all TB (CXR) | | | | | | | | | | |
| 1 | 5.0% (4.7-5.3%) | 10.1% (9.5-10.6%) | 15.2% (14.3-16.0%) | 20.3% (19.2-21.4%) | 25.4% (24.0-26.8%) | 30.6% (28.8-32.3%) | 35.9% (33.8-37.9%) | 41.1% (38.9-43.5%) | 46.5% (43.9-49.1%) | 51.8% (48.9-54.7%) |
| 2 | 9.3% (8.8-9.8%) | 18.1% (17.1-19.0%) | 26.5% (25.2-27.9%) | 34.3% (32.7-36.1%) | 41.8% (39.8-43.8%) | 48.6% (46.4-50.7%) | 55.0% (52.7-57.2%) | 60.8% (58.3-63.0%) | 66.1% (63.6-68.2%) | 70.7% (68.3-73.0%) |
| 3 | 12.9% (12.2-13.6%) | 24.5% (23.3-25.7%) | 34.9% (33.3-36.4%) | 44.0% (42.2-45.8%) | 52.1% (50.1-54.1%) | 59.1% (57.0-61.1%) | 65.0% (63.0-66.8%) | 69.9% (68.0-71.7%) | 74.0% (72.2-75.6%) | 77.2% (75.6-78.7%) |
| 4 | 15.9% (15.0-16.7%) | 29.4% (28.1-30.7%) | 40.9% (39.3-42.7%) | 50.6% (48.6-52.4%) | 58.3% (56.6-60.3%) | 64.8% (62.9-66.6%) | 69.9% (68.2-71.5%) | 73.9% (72.4-75.3%) | 77.1% (75.7-78.3%) | 79.5% (78.4-80.4%) |
| 5 | 18.2% (17.4-19.1%) | 33.2% (31.7-34.5%) | 45.1% (43.4-46.9%) | 54.8% (52.9-56.6%) | 62.2% (60.5-63.9%) | 67.9% (66.3-69.4%) | 72.4% (70.9-73.7%) | 75.7% (74.6-76.8%) | 78.2% (77.2-79.2%) | 80.2% (79.3-80.9%) |

Supplemental Table 12: Projected percentage of TB-associated deaths averted (2025-2035) by diagnostic algorithm, population coverage, and duration for a population with infectious TB prevalence of 500 per 100,000.

| | Projected reduction in TB-associated deaths (2025-2035) | | | | | | | | | |
|---|---|---|---|---|---|---|---|---|---|---|
| | Population coverage | | | | | | | | | |
| Rounds | 10% | 20% | 30% | 40% | 50% | 60% | 70% | 80% | 90% | 100% |
| Algorithm targeting symptomatic infectious TB (Cough+Xpert) | | | | | | | | | | |
| 1 | 1.0% (0.9-1.2%) | 2.0% (1.8-2.3%) | 3.0% (2.6-3.5%) | 4.0% (3.5-4.6%) | 5.0% (4.4-5.8%) | 6.0% (5.3-7.0%) | 7.0% (6.1-8.1%) | 8.0% (7.0-9.2%) | 9.1% (7.8-10.5%) | 10.1% (8.7-11.6%) |
| 2 | 1.9% (1.7-2.2%) | 3.8% (3.3-4.3%) | 5.7% (4.9-6.5%) | 7.4% (6.5-8.6%) | 9.2% (8.1-10.5%) | 10.9% (9.6-12.5%) | 12.6% (11.0-14.3%) | 14.1% (12.6-16.1%) | 15.7% (13.9-18%) | 17.3% (15.2-19.8%) |
| 3 | 2.8% (2.4-3.2%) | 5.5% (4.8-6.3%) | 8.1% (7.1-9.2%) | 10.5% (9.3-12.1%) | 13.0% (11.4-14.8%) | 15.2% (13.5-17.3%) | 17.4% (15.5-19.6%) | 19.6% (17.3-22.3%) | 21.6% (19.3-24.2%) | 23.6% (21.1-26.4%) |
| 4 | 3.6% (3.2-4.1%) | 7.0% (6.2-8.0%) | 10.3% (9.1-11.7%) | 13.4% (11.8-15.3%) | 16.4% (14.5-18.4%) | 19.1% (17.0-21.5%) | 21.8% (19.4-24.6%) | 24.3% (21.9-27.5%) | 26.8% (23.9-30.0%) | 29.2% (26.1-32.5%) |
| 5 | 4.3% (3.8-4.9%) | 8.4% (7.4-9.7%) | 12.3% (10.8-13.9%) | 15.9% (14.2-18.0%) | 19.4% (17.2-21.8%) | 22.6% (20.5-25.4%) | 25.8% (23.0-28.7%) | 28.8% (25.6-32.2%) | 31.5% (28.3-34.9%) | 34.1% (31.0-37.7%) |
| Algorithm targeting infectious TB (Xpert) | | | | | | | | | | |
| 1 | 2.4% (2.1-2.7%) | 4.7% (4.1-5.3%) | 7.1% (6.2-8.0%) | 9.4% (8.2-10.5%) | 11.8% (10.5-13.2%) | 14.2% (12.5-15.9%) | 16.5% (14.6-18.6%) | 19% (16.6-21.3%) | 21.2% (18.8-24.1%) | 23.6% (21-26.8%) |
| 2 | 4.5% (3.9-5.0%) | 8.8% (7.8-9.8%) | 12.9% (11.5-14.6%) | 16.9% (15.1-19.0%) | 20.8% (18.6-23.2%) | 24.5% (22-27.4%) | 28% (25-31.3%) | 31.5% (28.1-34.8%) | 34.7% (31.2-38.5%) | 37.8% (34.1-41.6%) |
| 3 | 6.4% (5.7-7.1%) | 12.3% (11.0-13.9%) | 17.9% (16.0-20.0%) | 23.3% (20.7-25.7%) | 28.2% (25.4-31.1%) | 32.8% (29.7-35.9%) | 37.1% (33.9-40.6%) | 41.1% (37.3-44.8%) | 44.8% (40.8-48.8%) | 48.3% (44.1-52.5%) |
| 4 | 8.0% (7.1-9.0%) | 15.4% (13.9-17.2%) | 22.1% (20.0-24.7%) | 28.4% (25.8-31.4%) | 34.2% (30.9-37.4%) | 39.4% (35.9-43.2%) | 44.2% (40.7-47.9%) | 48.5% (44.3-52.5%) | 52.5% (48.6-56.5%) | 56.2% (52.1-60%) |
| 5 | 9.5% (8.5-10.6%) | 18.1% (16.3-20.1%) | 25.9% (23.3-28.6%) | 32.8% (29.8-35.9%) | 39.0% (35.8-42.5%) | 44.6% (41.1-48.3%) | 49.6% (45.7-53.7%) | 54.2% (50.2-58%) | 58.2% (54.2-62%) | 61.8% (58.1-65.5%) |
| Algorithm targeting all TB (CXR) | | | | | | | | | | |
| 1 | 5.2% (4.9-5.5%) | 10.4% (9.9-10.9%) | 15.7% (14.8-16.5%) | 21.0% (19.9-22.0%) | 26.2% (24.8-27.6%) | 31.5% (29.8-33.2%) | 37.0% (35.0-38.9%) | 42.3% (40.2-44.7%) | 47.8% (45.3-50.4%) | 53.2% (50.5-56.1%) |
| 2 | 9.6% (9.1-10.1%) | 18.7% (17.8-19.6%) | 27.3% (26.0-28.6%) | 35.3% (33.7-37.1%) | 43.0% (41.0-44.9%) | 49.9% (47.8-51.9%) | 56.4% (54.2-58.6%) | 62.3% (59.8-64.3%) | 67.5% (65.3-69.6%) | 72.2% (69.9-74.3%) |
| 3 | 13.3% (12.6-14%) | 25.3% (24.1-26.5%) | 35.9% (34.4-37.4%) | 45.3% (43.4-47.0%) | 53.4% (51.5-55.4%) | 60.6% (58.5-62.5%) | 66.5% (64.7-68.2%) | 71.4% (69.6-73.1%) | 75.5% (73.7-77.0%) | 78.7% (77.2-80.0%) |
| 4 | 16.4% (15.6-17.2%) | 30.4% (29.1-31.6%) | 42.1% (40.5-43.8%) | 52.0% (50.0-53.7%) | 59.9% (58.1-61.6%) | 66.3% (64.5-68.0%) | 71.5% (69.9-72.9%) | 75.5% (74.0-76.8%) | 78.6% (77.4-79.7%) | 81.0% (80.0-81.9%) |
| 5 | 18.9% (18.0-19.8%) | 34.3% (32.8-35.5%) | 46.5% (44.8-48.2%) | 56.2% (54.4-57.9%) | 63.7% (62.2-65.3%) | 69.5% (67.9-70.9%) | 73.9% (72.6-75.1%) | 77.3% (76.2-78.3%) | 79.7% (78.7-80.6%) | 81.7% (80.8-82.4%) |

Supplemental Table 13: Projected reduction in TB mortality in 2035 by diagnostic algorithm, population coverage, and duration for a population with infectious prevalence of 500 per 100,000.

| | Projected reduction in 2035 TB mortality | | | | | | | | | |
|---|---|---|---|---|---|---|---|---|---|---|
| | Population coverage | | | | | | | | | |
| Rounds | 10% | 20% | 30% | 40% | 50% | 60% | 70% | 80% | 90% | 100% |
| Algorithm targeting symptomatic infectious TB (Cough+Xpert) | | | | | | | | | | |
| 1 | 0.7% (0.6-0.8%) | 1.4% (1.1-1.6%) | 2.1% (1.7-2.4%) | 2.8% (2.3-3.3%) | 3.4% (2.9-4.1%) | 4.1% (3.5-4.9%) | 4.8% (4.1-5.8%) | 5.5% (4.7-6.5%) | 6.2% (5.3-7.3%) | 6.9% (5.8-8.2%) |
| 2 | 1.4% (1.1-1.6%) | 2.7% (2.3-3.2%) | 4.0% (3.3-4.7%) | 5.3% (4.4-6.2%) | 6.6% (5.5-7.8%) | 7.8% (6.6-9.1%) | 8.9% (7.4-10.4%) | 10.1% (8.5-11.9%) | 11.2% (9.4-13.1%) | 12.3% (10.4-14.3%) |
| 3 | 2.1% (1.7-2.4%) | 4.0% (3.3-4.7%) | 6.0% (5.0-6.9%) | 7.7% (6.6-9.0%) | 9.5% (8.0-11.1%) | 11.2% (9.4-13.1%) | 12.8% (10.8-15.0%) | 14.4% (12.2-16.7%) | 15.9% (13.4-18.3%) | 17.3% (14.6-20.1%) |
| 4 | 2.7% (2.3-3.2%) | 5.3% (4.5-6.2%) | 7.8% (6.5-9.1%) | 10.2% (8.5-11.9%) | 12.4% (10.5-14.4%) | 14.5% (12.4-16.8%) | 16.6% (14-19.2%) | 18.5% (15.6-21.6%) | 20.4% (17.3-23.6%) | 22.1% (18.9-25.7%) |
| 5 | 3.4% (2.8-4.0%) | 6.6% (5.6-7.7%) | 9.7% (8.1-11.3%) | 12.6% (10.7-14.7%) | 15.3% (12.9-17.8%) | 17.9% (15.0-21.0%) | 20.2% (16.9-23.4%) | 22.4% (19.0-25.7%) | 24.7% (20.9-28.5%) | 26.7% (22.8-30.6%) |
| Algorithm targeting infectious TB (Xpert) | | | | | | | | | | |
| 1 | 1.8% (1.5-2.1%) | 3.7% (3.1-4.3%) | 5.5% (4.7-6.4%) | 7.4% (6.2-8.5%) | 9.3% (7.9-10.7%) | 11.2% (9.5-13.0%) | 13.0% (11.1-15.1%) | 15.0% (12.6-17.5%) | 16.9% (14.2-19.8%) | 18.8% (15.9-22.0%) |
| 2 | 3.6% (3.1-4.2%) | 7.2% (6.2-8.3%) | 10.7% (9.1-12.4%) | 14.0% (12.0-16.3%) | 17.4% (14.9-20.1%) | 20.5% (17.7-23.7%) | 23.6% (20.3-27.2%) | 26.7% (22.8-30.4%) | 29.5% (25.5-34.0%) | 32.3% (27.9-36.7%) |
| 3 | 5.5% (4.6-6.3%) | 10.7% (9.1-12.5%) | 15.6% (13.4-18.1%) | 20.5% (17.5-23.3%) | 24.9% (21.4-28.6%) | 29.1% (25.2-33.0%) | 33.2% (28.9-37.4%) | 36.9% (32.0-41.8%) | 40.5% (34.9-45.8%) | 43.7% (38.2-49.2%) |
| 4 | 7.3% (6.2-8.4%) | 14.1% (12.1-16.1%) | 20.4% (17.6-23.5%) | 26.3% (22.8-30.1%) | 31.8% (27.5-36.1%) | 36.9% (32.2-41.9%) | 41.6% (36.8-46.7%) | 45.8% (39.8-51.3%) | 49.9% (44.3-55.5%) | 53.7% (47.7-59.1%) |
| 5 | 9.1% (7.7-10.5%) | 17.4% (15.1-20.0%) | 25.0% (21.5-28.8%) | 32.0% (27.7-36.1%) | 38.2% (33.5-43.1%) | 43.9% (38.8-49.2%) | 49.0% (43.1-54.9%) | 53.7% (47.8-59.5%) | 57.9% (51.9-63.6%) | 61.8% (55.8-67.6%) |
| Algorithm targeting all TB (CXR) | | | | | | | | | | |
| 1 | 5.6% (5.3-5.9%) | 11.3% (10.6-11.9%) | 17% (16.1-18%) | 22.7% (21.5-24.1%) | 28.6% (27.0-30.3%) | 34.4% (32.6-36.5%) | 40.4% (38.2-42.8%) | 46.6% (43.9-49.2%) | 52.6% (49.5-55.6%) | 58.8% (55.6-62.2%) |
| 2 | 11.0% (10.3-11.6%) | 21.4% (20.3-22.6%) | 31.4% (29.8-33.0%) | 40.7% (38.8-42.8%) | 49.5% (47.1-51.8%) | 57.8% (55.1-60.2%) | 65.1% (62.5-67.8%) | 72.1% (69.1-74.7%) | 78.0% (75.4-80.9%) | 83.5% (80.6-86.1%) |
| 3 | 16.1% (15.3-17%) | 30.6% (29.2-32.1%) | 43.5% (41.5-45.5%) | 54.8% (52.5-57.2%) | 64.7% (62.1-67.0%) | 73.0% (70.5-75.4%) | 79.9% (77.5-82.2%) | 85.6% (83.4-87.9%) | 90.2% (88.1-91.9%) | 93.4% (91.8-95.0%) |
| 4 | 21.0% (19.8-22.1%) | 38.9% (37-40.7%) | 53.8% (51.5-55.9%) | 65.9% (63.4-68.2%) | 75.3% (73.1-77.7%) | 83% (80.8-84.9%) | 88.5% (86.5-90.2%) | 92.6% (91.1-94.0%) | 95.5% (94.3-96.5%) | 97.4% (96.5-98.2%) |
| 5 | 25.7% (24.4-27%) | 46.3% (44.2-48.3%) | 62.1% (60.0-64.3%) | 74.1% (72.0-76.4%) | 83% (80.9-84.9%) | 89.3% (87.5-90.8%) | 93.4% (92.1-94.7%) | 96.2% (95.2-97.1%) | 98.0% (97.3-98.5%) | 99.0% (98.5-99.3%) |

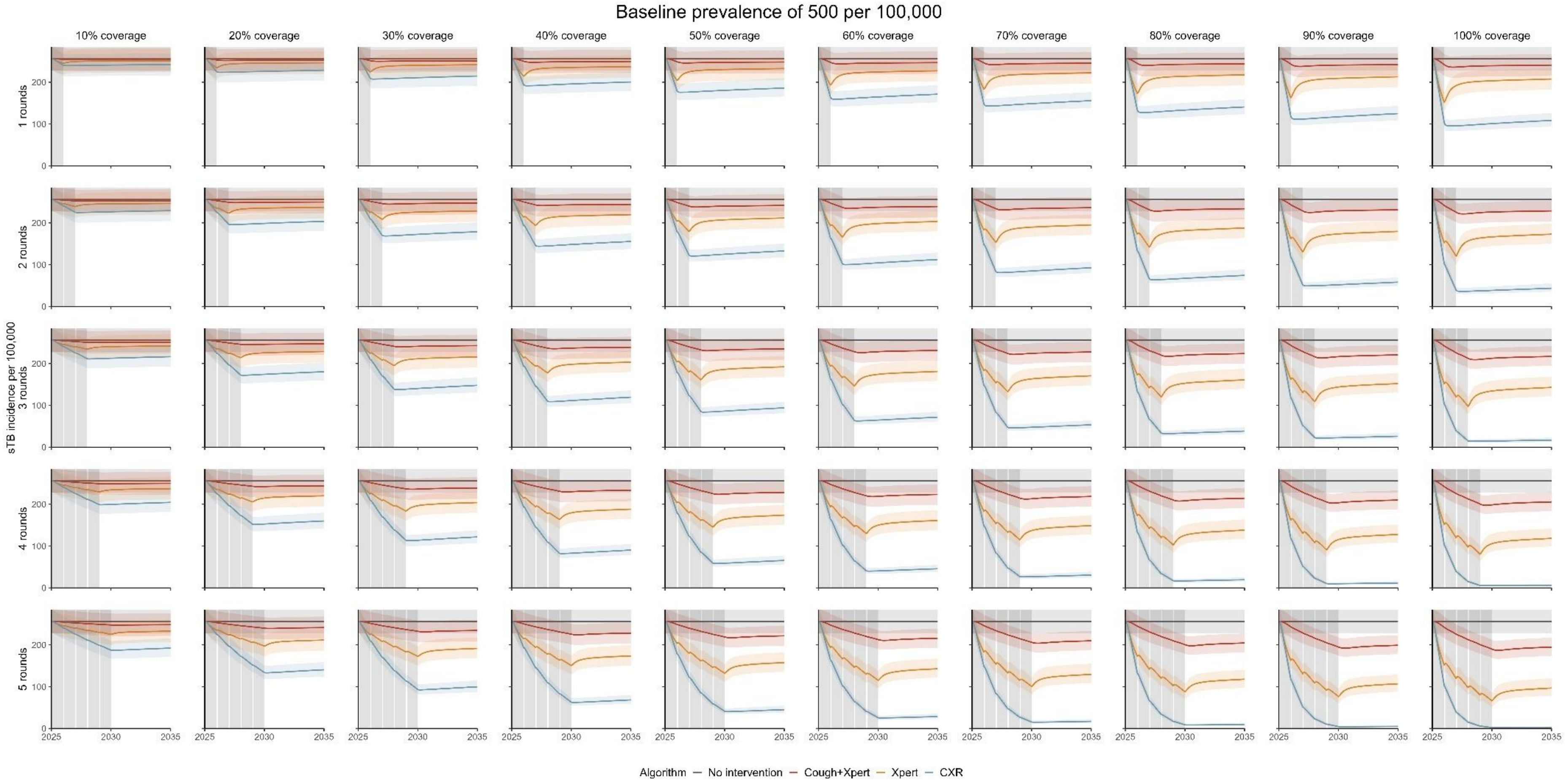


Supplemental Figure 4: Projected incidence of symptomatic TB by diagnostic algorithm, population coverage, and duration for a population with baseline prevalence of 500 per 100,000.

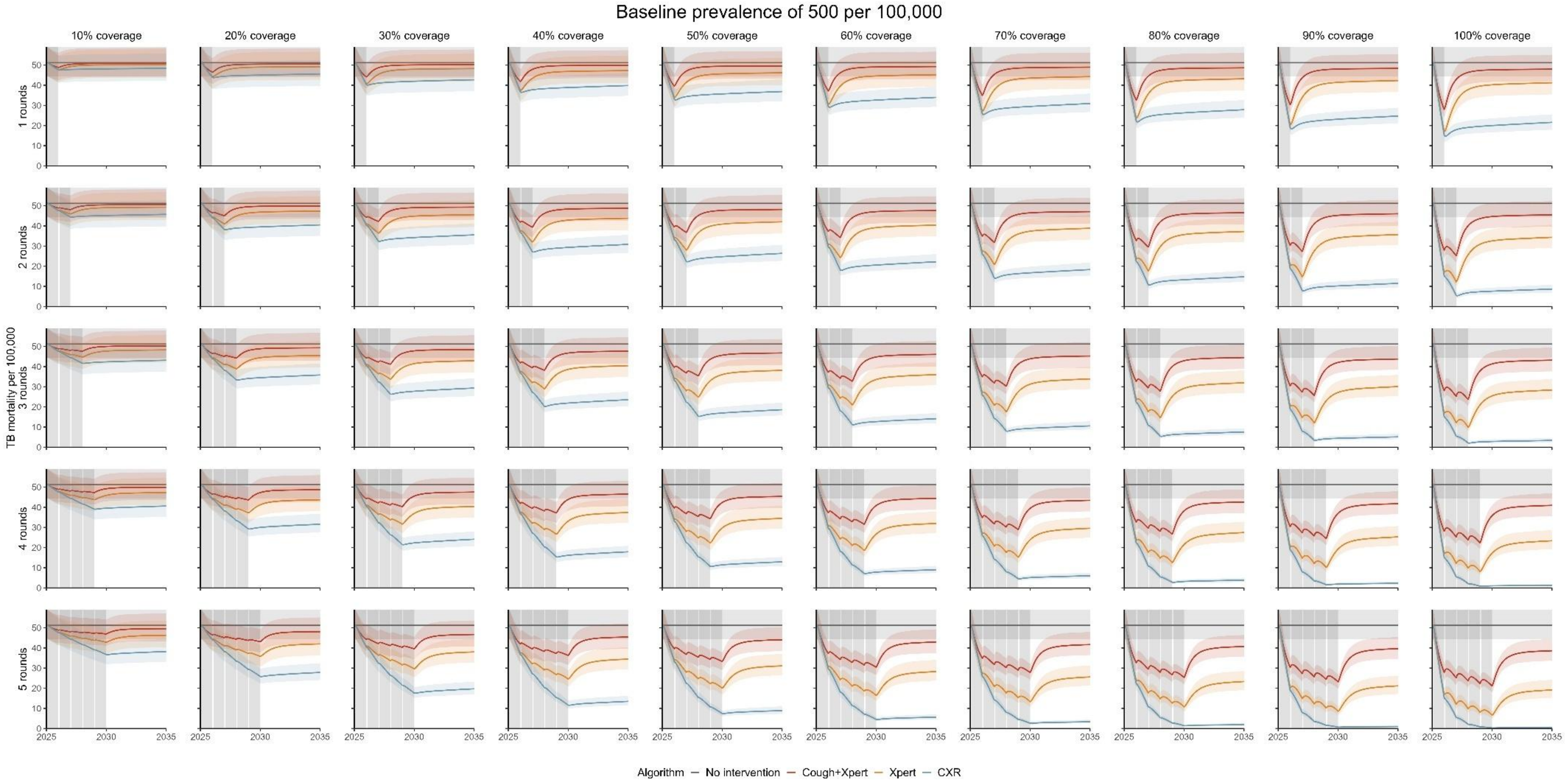


Supplemental Figure 5: Projected TB mortality by diagnostic algorithm, population coverage, and duration for a population with baseline prevalence of 500 per 100,000.

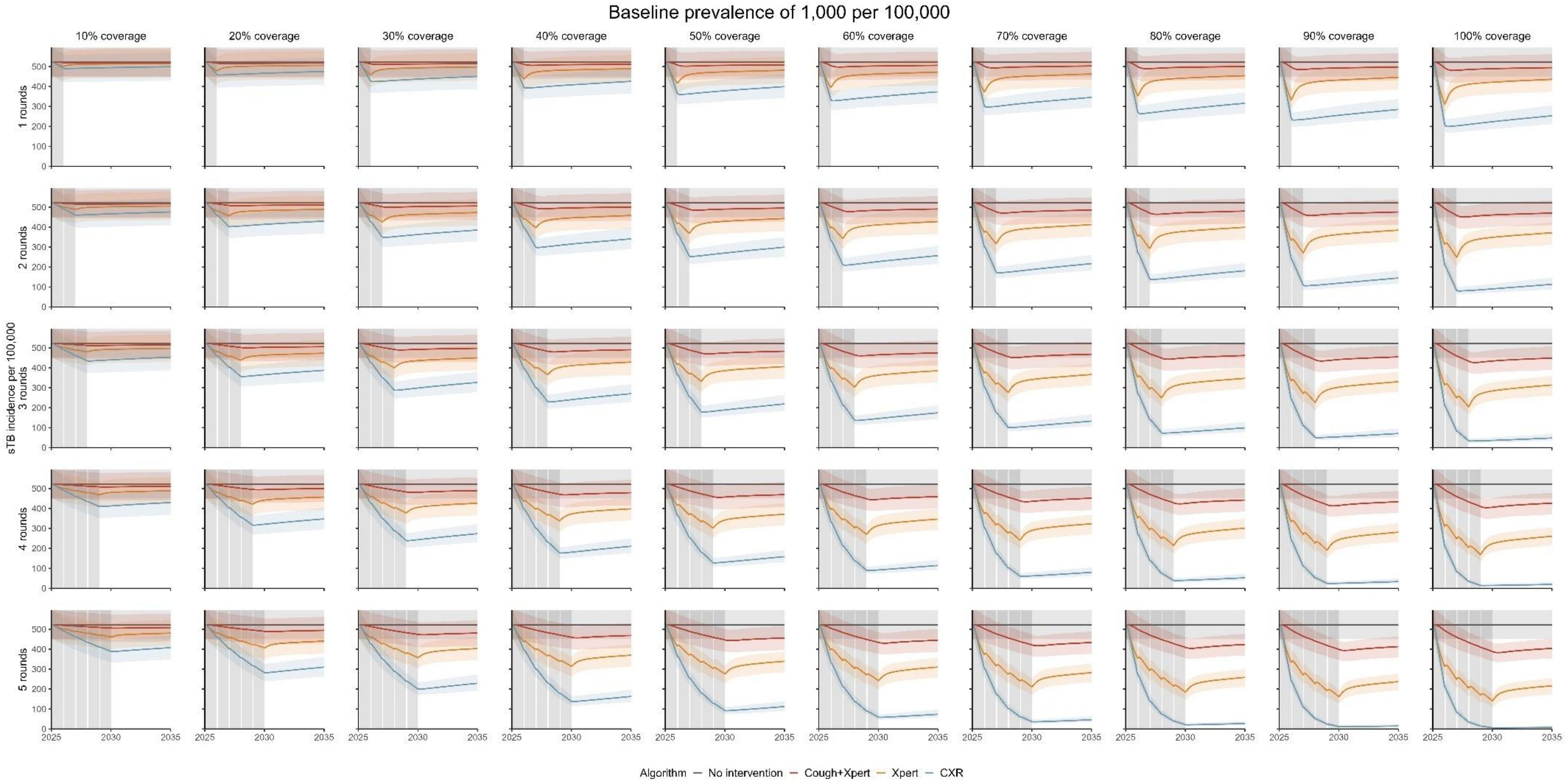


Supplemental Figure 6: Projected incidence of symptomatic TB by diagnostic algorithm, population coverage, and duration for a population with baseline prevalence of 1,000 per 100,000.

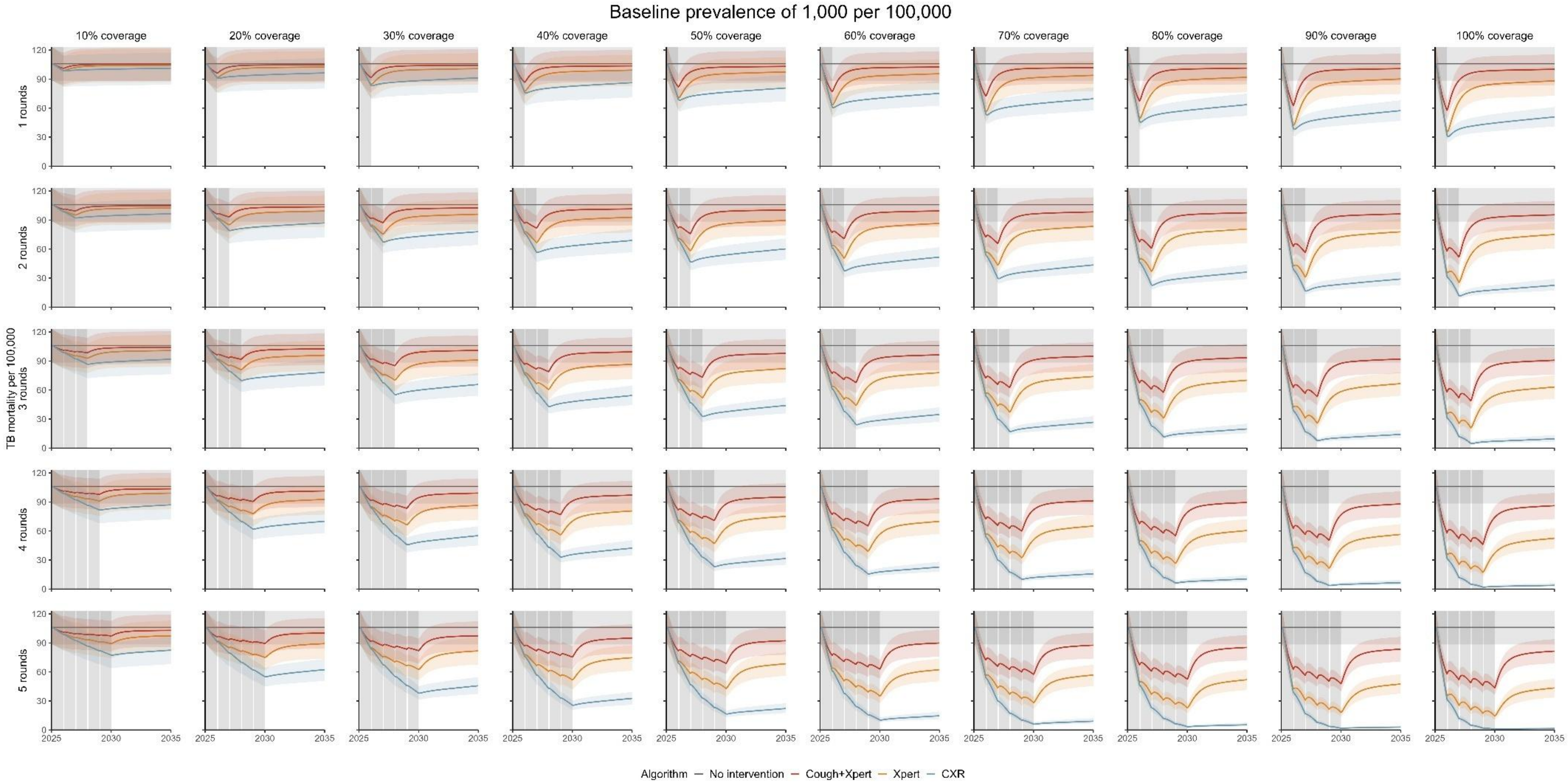


Supplemental Figure 7: Projected TB mortality by diagnostic algorithm, population coverage, and duration for a population with baseline prevalence of 1,000 per 100,000.

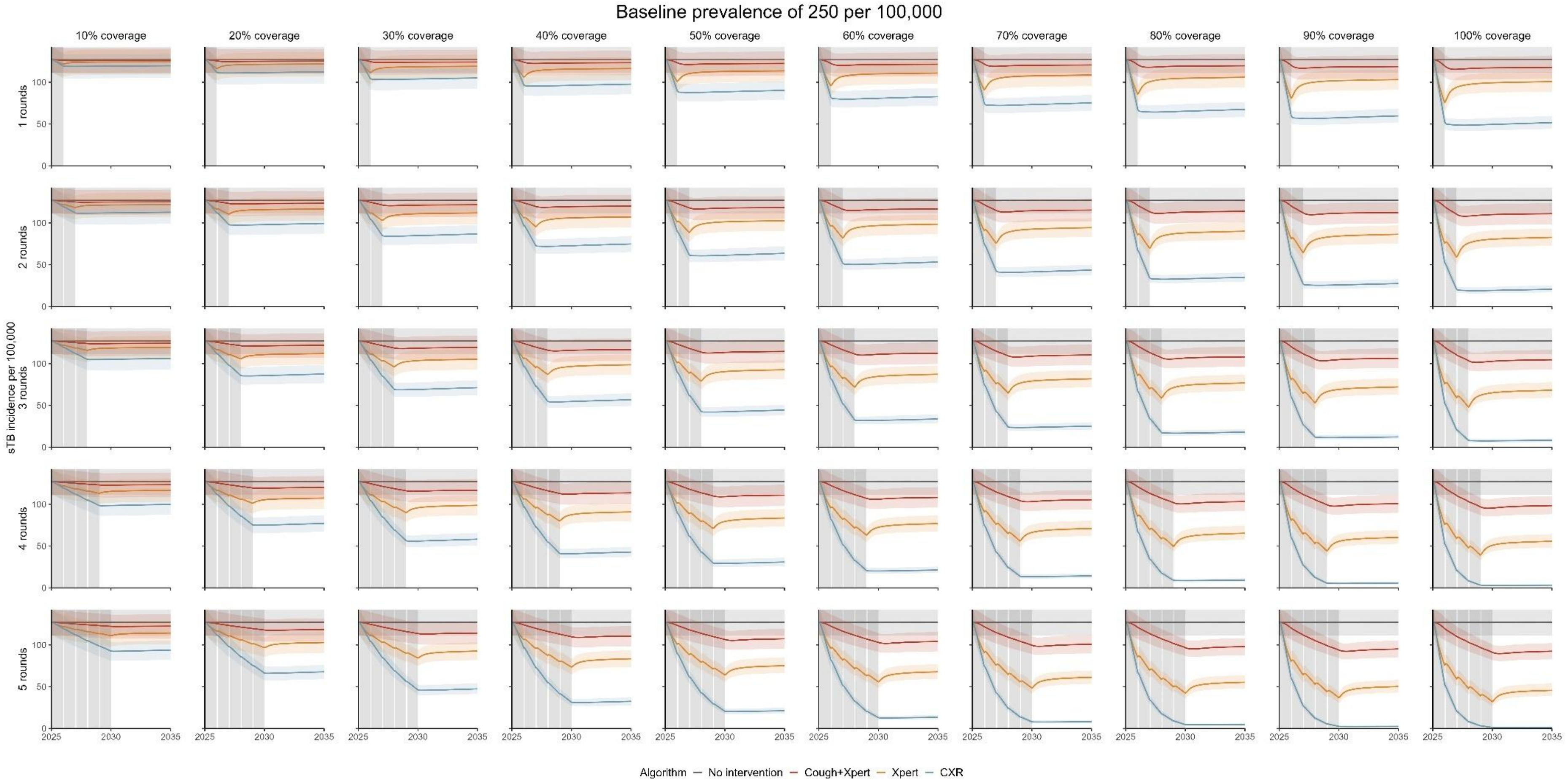


Supplemental Figure 8: Projected incidence of symptomatic TB by diagnostic algorithm, population coverage, and duration for a population with baseline prevalence of 250 per 100,000.

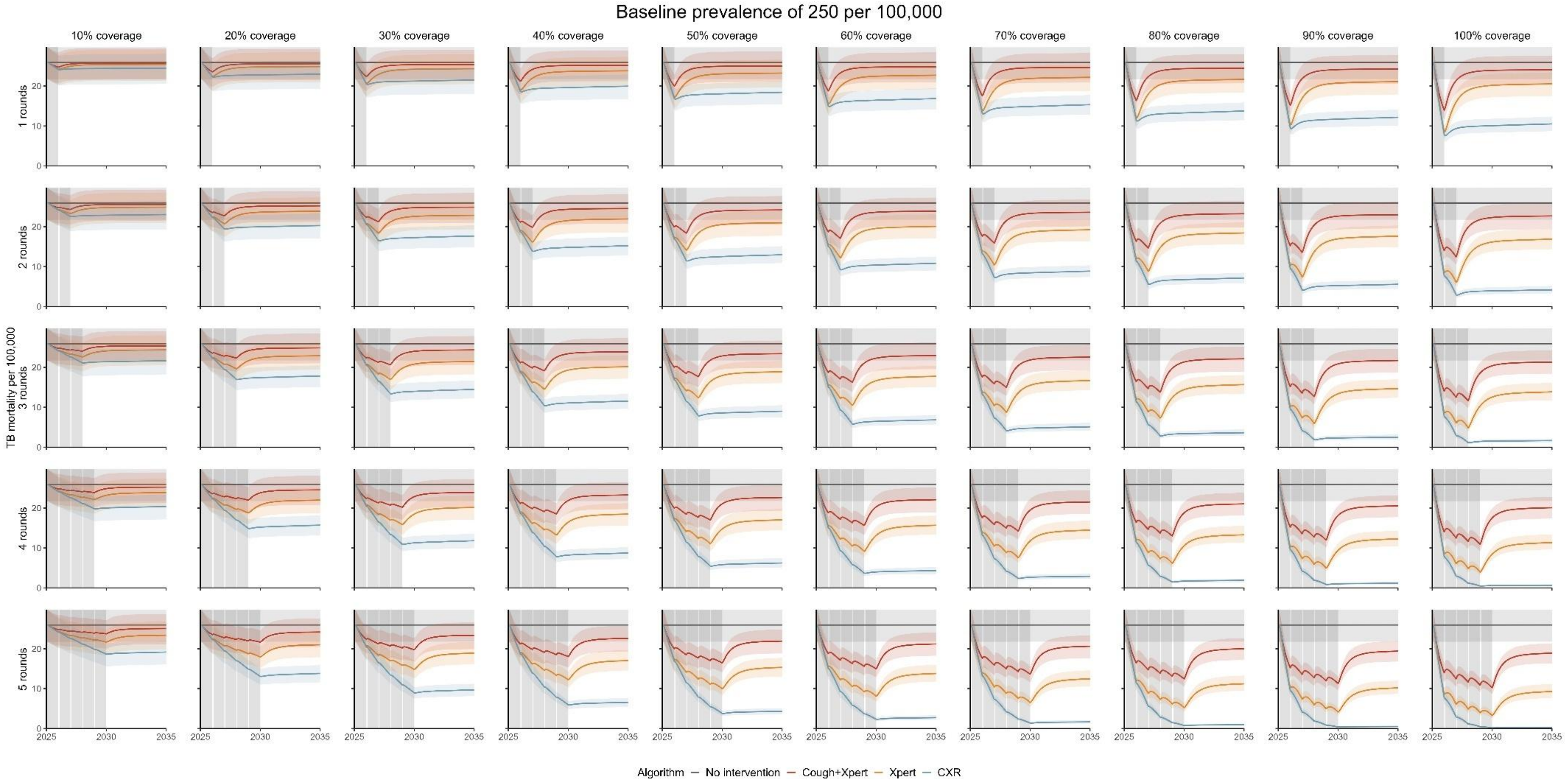


Supplemental Figure 9: Projected TB mortality by diagnostic algorithm, population coverage, and duration for a population with baseline prevalence of 250 per 100,000.

Supplemental Table 14: Projected reduction in symptomatic TB incidence at the end of screening by diagnostic algorithm, population coverage, and duration for baseline prevalence of 500 per 100,000 population.

| | Projected reduction in symptomatic TB incidence at the end of screening | | | | | | | | | |
|---|---|---|---|---|---|---|---|---|---|---|
| | Population coverage | | | | | | | | | |
| Rounds | 10% | 20% | 30% | 40% | 50% | 60% | 70% | 80% | 90% | 100% |
| Algorithm targeting symptomatic infectious TB (Cough+Xpert) | | | | | | | | | | |
| 1 | 0.8% (0.7-1.0%) | 1.6% (1.3-1.9%) | 2.4% (2.0-2.9%) | 3.2% (2.6-3.9%) | 4.0% (3.2-4.8%) | 4.8% (3.9-5.8%) | 5.6% (4.6-6.7%) | 6.4% (5.2-7.7%) | 7.2% (5.9-8.7%) | 7.9% (6.5-9.7%) |
| 2 | 1.5% (1.2-1.8%) | 3.0% (2.5-3.6%) | 4.4% (3.7-5.4%) | 5.8% (4.8-7.1%) | 7.2% (6.0-8.7%) | 8.6% (7.1-10.3%) | 9.8% (8.1-11.8%) | 11.1% (9.2-13.3%) | 12.3% (10.2-14.9%) | 13.5% (11.1-16.2%) |
| 3 | 2.2% (1.8-2.6%) | 4.3% (3.5-5.1%) | 6.3% (5.2-7.5%) | 8.2% (6.8-10.0%) | 10.1% (8.3-12.0%) | 11.8% (9.9-14.1%) | 13.5% (11.3-16.0%) | 15.2% (12.7-18.2%) | 16.8% (14.1-19.9%) | 18.3% (15.4-21.8%) |
| 4 | 2.8% (2.3-3.4%) | 5.5% (4.6-6.6%) | 8.0% (6.7-9.7%) | 10.5% (8.7-12.5%) | 12.9% (10.7-15.3%) | 15.0% (12.6-17.7%) | 17.1% (14.3-20.3%) | 19.0% (15.9-22.5%) | 20.9% (17.6-24.7%) | 22.7% (19.0-26.8%) |
| 5 | 3.4% (2.8-4.2%) | 6.7% (5.4-8.1%) | 9.7% (8.0-11.6%) | 12.6% (10.5-15.0%) | 15.3% (12.8-18.2%) | 17.8% (15.2-21.2%) | 20.3% (17.2-23.9%) | 22.7% (18.9-26.9%) | 24.8% (21.0-29.2%) | 26.9% (22.8-31.5%) |
| Algorithm targeting infectious TB (Xpert) | | | | | | | | | | |
| 1 | 4.1% (3.6-4.6%) | 8.2% (7.2-9.2%) | 12.3% (10.7-13.8%) | 16.2% (14.3-18.4%) | 20.4% (17.8-22.9%) | 24.5% (21.5-27.4%) | 28.3% (25.2-32.0%) | 32.7% (28.5-36.6%) | 36.4% (32.2-41.2%) | 40.5% (35.8-45.5%) |
| 2 | 6.5% (5.6-7.4%) | 12.7% (11.1-14.4%) | 18.7% (16.4-21.1%) | 24.3% (21.5-27.5%) | 29.8% (26.3-33.6%) | 34.9% (30.9-39.3%) | 39.7% (35.2-44.8%) | 44.5% (39.5-49.5%) | 48.9% (43.5-54.3%) | 52.9% (47.6-58.7%) |
| 3 | 8.5% (7.4-9.7%) | 16.4% (14.3-18.8%) | 23.8% (20.8-26.9%) | 30.6% (26.8-34.4%) | 37.0% (32.7-41.3%) | 42.8% (37.8-47.5%) | 48.2% (43.2-53.2%) | 53.1% (47.4-58.6%) | 57.5% (51.5-63.1%) | 61.6% (55.6-67.4%) |
| 4 | 10.4% (9-11.9%) | 19.8% (17.3-22.4%) | 28.2% (24.9-32.1%) | 36.0% (32.0-40.5%) | 43.0% (38.1-48.2%) | 49.2% (44.0-54.6%) | 55.0% (49.8-60.3%) | 59.8% (53.9-65.5%) | 64.6% (58.8-69.8%) | 68.7% (62.7-73.9%) |
| 5 | 12.2% (10.6-13.9%) | 23.0% (20.2-26%) | 32.6% (28.5-36.9%) | 41.1% (36.3-45.6%) | 48.5% (43.2-53.7%) | 54.9% (49.6-60.1%) | 60.6% (54.7-66.4%) | 65.7% (59.9-71.1%) | 70.1% (64.2-75.4%) | 74.0% (68.5-79.1%) |
| Algorithm targeting all TB (CXR) | | | | | | | | | | |
| 1 | 6.3% (6.0-6.6%) | 12.7% (12.0-13.3%) | 18.9% (18.0-19.9%) | 25.2% (24-26.5%) | 31.4% (29.9-33.0%) | 37.7% (35.8-39.7%) | 44% (41.7-46.2%) | 50.1% (47.7-52.7%) | 56.4% (53.6-59.4%) | 62.6% (59.4-65.8%) |
| 2 | 12.2% (11.6-12.8%) | 23.5% (22.4-24.6%) | 34.1% (32.5-35.6%) | 43.8% (42.0-45.8%) | 52.9% (50.7-55.0%) | 60.9% (58.5-63.2%) | 68.3% (65.7-70.8%) | 75.0% (72.2-77.3%) | 80.7% (78.3-83.0%) | 85.7% (83.1-88.1%) |
| 3 | 17.5% (16.6-18.4%) | 32.9% (31.5-34.4%) | 46.2% (44.3-48.1%) | 57.6% (55.4-59.7%) | 67.3% (65.0-69.6%) | 75.5% (73.1-77.7%) | 82.0% (79.9-84.0%) | 87.2% (85.3-89.1%) | 91.4% (89.6-93.0%) | 94.4% (92.9-95.8%) |
| 4 | 22.5% (21.4-23.6%) | 40.9% (39.2-42.6%) | 55.9% (53.9-58.1%) | 67.9% (65.6-70.1%) | 77.2% (75.1-79.3%) | 84.4% (82.4-86.3%) | 89.7% (88.0-91.2%) | 93.5% (92.1-94.7%) | 96.1% (95.0-97.1%) | 97.9% (97.1-98.5%) |
| 5 | 27.0% (25.8-28.2%) | 48.0% (45.9-49.8%) | 63.8% (61.6-66.0%) | 75.7% (73.4-77.8%) | 84.1% (82.3-86%) | 90.1% (88.4-91.5%) | 94.1% (92.8-95.2%) | 96.7% (95.8-97.5%) | 98.3% (97.6-98.8%) | 99.2% (98.8-99.4%) |

Supplemental Table 15: Projected reduction in TB mortality at the end of screening by diagnostic algorithm, population coverage, and duration for baseline prevalence of 500 per 100,000 population.

| | Projected reduction in TB mortality at the end of screening | | | | | | | | | |
|---|---|---|---|---|---|---|---|---|---|---|
| | Population coverage | | | | | | | | | |
| Rounds | 10% | 20% | 30% | 40% | 50% | 60% | 70% | 80% | 90% | 100% |
| Algorithm targeting symptomatic infectious TB (Cough+Xpert) | | | | | | | | | | |
| 1 | 4.5% (4.1-4.9%) | 9.0% (8.2-9.9%) | 13.6% (12.4-14.9%) | 18.2% (16.5-19.8%) | 22.6% (20.8-24.8%) | 27.1% (24.7-29.6%) | 31.6% (28.7-34.5%) | 36.2% (33.0-39.5%) | 40.8% (36.8-44.6%) | 45.1% (41.0-49.4%) |
| 2 | 6.0% (5.4-6.7%) | 11.8% (10.7-13.1%) | 17.4% (15.8-19.3%) | 22.9% (20.8-25.3%) | 28.0% (25.5-30.6%) | 32.9% (30.1-36.2%) | 37.7% (34.5-41.2%) | 42.2% (38.8-46.1%) | 46.5% (42.6-50.5%) | 50.8% (46.7-55.0%) |
| 3 | 6.8% (6.1-7.7%) | 13.4% (12.0-14.8%) | 19.5% (17.6-21.5%) | 25.3% (22.8-28.1%) | 30.7% (27.9-33.8%) | 35.8% (32.6-39.2%) | 40.6% (37.1-44.2%) | 45.3% (41.4-49.2%) | 49.6% (45.7-53.7%) | 53.7% (49.8-57.7%) |
| 4 | 7.5% (6.7-8.5%) | 14.5% (13.0-16.3%) | 21.1% (19.0-23.5%) | 27.3% (24.6-30.1%) | 32.9% (30.0-36.3%) | 38.3% (34.7-41.7%) | 43.2% (39.6-47.2%) | 47.7% (44.0-52.0%) | 52.1% (48.0-56.5%) | 56.3% (51.9-60.7%) |
| 5 | 8.1% (7.2-9.2%) | 15.6% (13.9-17.6%) | 22.6% (20.2-25.2%) | 29.0% (26.2-32.1%) | 34.9% (31.6-38.5%) | 40.4% (37.1-44.4%) | 45.5% (41.6-49.6%) | 50.4% (45.8-54.9%) | 54.6% (50.4-59.1%) | 58.7% (54.5-63.2%) |
| Algorithm targeting infectious TB (Xpert) | | | | | | | | | | |
| 1 | 6.7% (6.3-7.1%) | 13.4% (12.6-14.2%) | 20.1% (19.0-21.3%) | 26.7% (25.3-28.2%) | 33.4% (31.5-35.3%) | 40.2% (37.9-42.5%) | 46.8% (44.3-49.4%) | 53.5% (50.4-56.6%) | 60.1% (56.8-63.4%) | 66.6% (63.1-70.4%) |
| 2 | 10.2% (9.5-11.0%) | 19.8% (18.5-21.2%) | 28.9% (27.0-30.9%) | 37.3% (35.0-39.9%) | 45.2% (42.4-47.8%) | 52.4% (49.5-55.6%) | 59.2% (55.7-62.8%) | 65.5% (62.1-68.9%) | 71.1% (67.6-74.6%) | 76.1% (72.6-79.7%) |
| 3 | 12.5% (11.5-13.5%) | 23.8% (21.9-25.9%) | 34.0% (31.5-36.7%) | 43.3% (40.2-46.2%) | 51.6% (48.2-54.9%) | 58.9% (55.4-62.4%) | 65.4% (62.0-69.1%) | 71.2% (67.3-74.7%) | 76.2% (72.4-79.6%) | 80.7% (77.3-84.0%) |
| 4 | 14.4% (13.1-15.6%) | 27.1% (25.0-29.4%) | 38.1% (35.3-41.4%) | 47.8% (44.5-51.4%) | 56.3% (52.4-60.1%) | 63.7% (59.9-67.7%) | 70.1% (66.3-74.0%) | 75.4% (71.4-79.1%) | 80.1% (76.8-83.5%) | 84.2% (80.7-87.1%) |
| 5 | 16.1% (14.7-17.7%) | 29.9% (27.4-32.7%) | 41.9% (38.4-45.3%) | 52.0% (48.1-55.7%) | 60.5% (56.7-64.6%) | 67.7% (63.8-71.6%) | 73.8% (69.8-77.8%) | 79.0% (75.2-82.5%) | 83.3% (79.7-86.4%) | 86.9% (83.9-89.7%) |
| Algorithm targeting all TB (CXR) | | | | | | | | | | |
| 1 | 7.2% (6.8-7.6%) | 14.4% (13.7-15.1%) | 21.5% (20.5-22.7%) | 28.7% (27.3-30.1%) | 35.9% (34.0-37.8%) | 43.1% (40.8-45.2%) | 50.0% (47.6-52.8%) | 57.3% (54.4-60.4%) | 64.2% (61.1-67.9%) | 71.4% (67.8-75.3%) |
| 2 | 13.3% (12.6-13.9%) | 25.5% (24.4-26.6%) | 36.8% (35.3-38.4%) | 47.1% (45.2-49.1%) | 56.6% (54.5-58.8%) | 65.0% (62.6-67.3%) | 72.6% (70.1-75.0%) | 79.2% (76.7-81.4%) | 84.8% (82.6-87.0%) | 89.5% (87.4-91.5%) |
| 3 | 18.6% (17.8-19.4%) | 34.8% (33.3-36.2%) | 48.6% (46.9-50.4%) | 60.4% (58.2-62.3%) | 70.2% (68.0-72.2%) | 78.2% (76.0-80.2%) | 84.6% (82.7-86.3%) | 89.5% (87.8-91.2%) | 93.3% (91.7-94.6%) | 96.0% (94.8-97.0%) |
| 4 | 23.6% (22.5-24.7%) | 42.7% (41.1-44.3%) | 58% (56.1-60.1%) | 70.1% (67.9-72.1%) | 79.2% (77.3-81.1%) | 86.2% (84.4-87.8%) | 91.2% (89.7-92.5%) | 94.7% (93.4-95.7%) | 97.0% (96.1-97.8%) | 98.5% (97.9-98.9%) |
| 5 | 28.1% (26.9-29.3%) | 49.7% (47.8-51.3%) | 65.6% (63.6-67.6%) | 77.3% (75.3-79.3%) | 85.6% (83.9-87.2%) | 91.2% (89.7-92.5%) | 95% (93.9-95.9%) | 97.3% (96.6-97.9%) | 98.7% (98.1-99.0%) | 99.4% (99.1-99.6%) |

Supplemental Table 16: Rebound in sTB incidence between the end of screening and 10 years after the start of screening by diagnostic algorithm, population coverage, and duration of screening

| | Rebound in sTB incidence between the end of screening and 10 years after the start of screening | | | | | | | | | |
|---|---|---|---|---|---|---|---|---|---|---|
| | Population coverage | | | | | | | | | |
| Rounds | 10% | 20% | 30% | 40% | 50% | 60% | 70% | 80% | 90% | 100% |
| Symptomatic infectious TB (Cough+Xpert) | | | | | | | | | | |
| 1 | 25.3% (19.6-31.5%) | 25.1% (19.5-31.3%) | 25.1% (19.5-31.2%) | 25.0% (19.2-31.1%) | 24.9% (19.3-31.1%) | 24.9% (19.3-31.1%) | 24.7% (19.0-30.9%) | 24.7% (18.9-30.8%) | 24.5% (18.9-30.8%) | 24.5% (18.9-30.4%) |
| 2 | 21.5% (16.6-26.9%) | 21.2% (16.4-26.9%) | 21.0% (16.4-26.8%) | 20.9% (16.2-26.4%) | 20.7% (15.9-26.2%) | 20.5% (15.8-26.0%) | 20.4% (15.7-25.8%) | 20.2% (15.3-25.8%) | 20.0% (15.2-25.4%) | 19.8% (15.3-25.3%) |
| 3 | 17.7% (13.8-22.6%) | 17.5% (13.6-22.2%) | 17.2% (13.3-22.0%) | 17.1% (13.1-21.8%) | 16.8% (12.9-21.7%) | 16.6% (12.8-21.2%) | 16.4% (12.6-21.0%) | 16.2% (12.3-21.0%) | 16.0% (12.2-20.6%) | 15.7% (12.2-20.4%) |
| 4 | 14.6% (11.3-18.7%) | 14.3% (11.2-18.6%) | 14.0% (10.8-18.0%) | 13.8% (10.6-18.0%) | 13.6% (10.4-17.5%) | 13.4% (10.3-17.2%) | 13.1% (10.0-17.2%) | 12.9% (9.8-16.7%) | 12.7% (9.6-16.4%) | 12.5% (9.5-16.2%) |
| 5 | 11.9% (9.2-15.4%) | 11.6% (8.9-15.1%) | 11.3% (8.7-14.9%) | 11.1% (8.5-14.5%) | 10.9% (8.3-14.3%) | 10.6% (8.0-13.9%) | 10.4% (7.9-13.6%) | 10.2% (7.6-13.3%) | 10% (7.5-13.3%) | 9.8% (7.4-12.9%) |
| Infectious TB (Xpert) | | | | | | | | | | |
| 1 | 54.7% (50.7-58.3%) | 54.5% (50.7-58.4%) | 54.4% (50.3-58.0%) | 54% (49.8-57.9%) | 53.9% (49.8-57.9%) | 53.6% (49.4-57.6%) | 53.4% (49.3-57.7%) | 53.3% (49.2-57.2%) | 53% (49.1-57.2%) | 52.8% (48.6-57.0%) |
| 2 | 42.9% (39.5-46.2%) | 42.3% (38.9-45.8%) | 41.9% (38.4-45.5%) | 41.3% (38.1-44.8%) | 40.8% (37.5-44.5%) | 40.4% (37.0-43.9%) | 39.9% (36.4-43.6%) | 39.3% (35.9-43.0%) | 38.9% (35.5-42.7%) | 38.4% (35.1-41.9%) |
| 3 | 34.9% (31.8-38.1%) | 34.1% (31.0-37.1%) | 33.4% (30.1-36.5%) | 32.5% (29.7-35.5%) | 31.8% (28.9-35.0%) | 31% (27.9-34.2%) | 30.3% (27.4-33.7%) | 29.6% (26.6-33.0%) | 29.0% (26.0-32.5%) | 28.3% (25.0-31.6%) |
| 4 | 29.0% (26.3-31.7%) | 27.9% (25.3-30.9%) | 27.0% (24.4-29.9%) | 26.1% (23.4-28.8%) | 25.2% (22.5-28.1%) | 24.3% (21.6-27.2%) | 23.5% (20.6-26.4%) | 22.8% (20.0-25.7%) | 22.0% (19.3-24.9%) | 21.2% (18.5-24.0%) |
| 5 | 24.3% (22.1-26.8%) | 23.2% (20.8-25.8%) | 22.1% (19.7-24.6%) | 21.1% (18.8-23.7%) | 20.1% (17.8-22.8%) | 19.2% (16.9-21.8%) | 18.3% (16.0-21.0%) | 17.5% (15.1-20.2%) | 16.7% (14.4-19.3%) | 15.9% (13.5-18.6%) |
| All TB (CXR) | | | | | | | | | | |
| 1 | 15.5% (13.1-18.6%) | 14.8% (12.5-17.8%) | 14.0% (11.7-17.0%) | 13.2% (11-16.1%) | 12.5% (10.2-15.3%) | 11.5% (9.5-14.4%) | 10.7% (8.7-13.5%) | 9.8% (7.9-12.6%) | 8.9% (7.1-11.5%) | 7.9% (6.2-10.5%) |
| 2 | 13.4% (11.2-15.8%) | 12.1% (10.2-14.5%) | 10.9% (9.0-13.2%) | 9.6% (8.0-11.9%) | 8.4% (6.8-10.6%) | 7.4% (5.8-9.2%) | 6.3% (4.9-8.0%) | 5.2% (3.9-6.8%) | 4.1% (3.1-5.6%) | 3.2% (2.3-4.6%) |
| 3 | 11.2% (9.3-13.2%) | 9.7% (8.0-11.5%) | 8.1% (6.7-9.9%) | 6.8% (5.5-8.3%) | 5.5% (4.3-7.0%) | 4.3% (3.3-5.7%) | 3.4% (2.5-4.5%) | 2.5% (1.8-3.4%) | 1.7% (1.2-2.5%) | 1.1% (0.7-1.8%) |
| 4 | 9.1% (7.6-10.8%) | 7.4% (6.1-8.9%) | 5.9% (4.7-7.2%) | 4.5% (3.6-5.7%) | 3.4% (2.6-4.4%) | 2.5% (1.8-3.3%) | 1.7% (1.2-2.4%) | 1.1% (0.7-1.6%) | 0.6% (0.4-1.0%) | 0.4% (0.2-0.6%) |
| 5 | 7.1% (5.8-8.6%) | 5.4% (4.4-6.6%) | 4% (3.2-5.0%) | 2.9% (2.2-3.7%) | 2.0% (1.4-2.7%) | 1.3% (0.9-1.8%) | 0.8% (0.5-1.2%) | 0.4% (0.3-0.7%) | 0.2% (0.1-0.4%) | 0.1% (0.1-0.2%) |

Supplemental Table 17: Rebound in TB mortality between the end of screening and 10 years after the start of screening by diagnostic algorithm, population coverage, and duration of screening

| | Rebound in TB mortality between the end of screening and 10 years after the start of screening | | | | | | | | | |
|---|---|---|---|---|---|---|---|---|---|---|
| | Population coverage | | | | | | | | | |
| Rounds | 10% | 20% | 30% | 40% | 50% | 60% | 70% | 80% | 90% | 100% |
| Symptomatic infectious TB (Cough+Xpert) | | | | | | | | | | |
| 1 | 86.6% (84.4-88.6%) | 86.7% (84.5-88.6%) | 86.7% (84.4-88.5%) | 86.6% (84.4-88.6%) | 86.6% (84.3-88.5%) | 86.6% (84.4-88.5%) | 86.5% (84.3-88.5%) | 86.5% (84.2-88.5%) | 86.5% (84.2-88.4%) | 86.5% (84.2-88.5%) |
| 2 | 80.1% (77.2-82.6%) | 79.9% (76.9-82.5%) | 79.7% (76.7-82.4%) | 79.5% (76.5-82.4%) | 79.4% (76.5-82%) | 79.2% (76.1-81.9%) | 79.0% (76.1-81.8%) | 78.9% (75.7-81.6%) | 78.7% (75.4-81.5%) | 78.4% (75.2-81.1%) |
| 3 | 73.6% (70.3-76.6%) | 73.4% (69.9-76.5%) | 73.0% (69.6-76.1%) | 72.8% (69.3-76.0%) | 72.5% (69.0-75.6%) | 72.2% (68.6-75.7%) | 71.8% (68.3-75.1%) | 71.5% (67.8-74.9%) | 71.3% (67.6-74.8%) | 71.0% (67.1-74.6%) |
| 4 | 67.8% (64.0-71.2%) | 67.4% (63.7-70.8%) | 66.9% (63.0-70.5%) | 66.5% (62.7-70.2%) | 66.1% (62.2-69.8%) | 65.8% (61.8-69.6%) | 65.4% (61.3-69.4%) | 64.9% (60.8-69.0%) | 64.7% (60.4-68.7%) | 64.3% (60.2-68.5%) |
| 5 | 62.4% (58.5-66.1%) | 61.8% (57.7-65.8%) | 61.5% (57.4-65.4%) | 60.9% (56.9-64.9%) | 60.5% (56.4-64.6%) | 60% (55.5-64.0%) | 59.7% (55.1-63.7%) | 59.1% (54.3-63.6%) | 58.6% (54.0-63.0%) | 58.2% (53.6-63.0%) |
| Infectious TB (Xpert) | | | | | | | | | | |
| 1 | 71.9% (68.1-75.5%) | 71.9% (68.4-75.5%) | 71.8% (67.9-75.4%) | 71.6% (68.2-75.4%) | 71.6% (68.0-75.2%) | 71.5% (67.8-75.1%) | 71.4% (67.8-75.0%) | 71.3% (67.6-75.1%) | 71.2% (67.4-75.1%) | 71.0% (67.4-74.8%) |
| 2 | 63.2% (59.5-67.0%) | 62.7% (58.7-66.5%) | 62.0% (58.1-66.0%) | 61.4% (57.5-65.4%) | 60.6% (56.6-65.1%) | 60.0% (56.0-64.2%) | 59.2% (55.1-63.4%) | 58.4% (54.3-62.9%) | 57.6% (53.3-61.8%) | 56.7% (52.3-61.3%) |
| 3 | 55.0% (51.3-59.1%) | 54.0% (50.0-58.1%) | 52.8% (49.0-57.4%) | 51.7% (47.9-56.1%) | 50.6% (46.3-55.0%) | 49.4% (45.5-54.0%) | 48.4% (44.4-52.8%) | 47.1% (42.8-51.9%) | 45.9% (41.5-51.0%) | 44.8% (40.1-49.9%) |
| 4 | 48% (44.4-51.7%) | 46.6% (42.7-50.8%) | 45.2% (41.2-49.5%) | 43.8% (39.7-48.0%) | 42.3% (38.3-46.8%) | 40.8% (36.6-45.5%) | 39.5% (35.4-44.0%) | 38.2% (33.6-43.2%) | 36.8% (32.4-41.6%) | 35.3% (30.9-40.2%) |
| 5 | 41.9% (38.4-45.9%) | 40.3% (36.9-44.3%) | 38.7% (34.6-43.3%) | 37.1% (33.3-41.5%) | 35.4% (31.5-39.9%) | 33.9% (29.8-38.3%) | 32.4% (28.2-37.2%) | 30.9% (26.6-35.7%) | 29.4% (25.1-34.2%) | 27.9% (23.6-32.8%) |
| All TB (CXR) | | | | | | | | | | |
| 1 | 24.7% (19.9-29.8%) | 24.1% (19.3-29.1%) | 23.4% (18.4-28.5%) | 22.7% (17.9-28.0%) | 22.5% (17.4-27.4%) | 21.8% (16.7-26.6%) | 20.8% (15.6-26.2%) | 20.3% (15.2-25.3%) | 19.4% (14.4-24.5%) | 18.8% (13.7-23.7%) |
| 2 | 19.6% (15.7-23.2%) | 18.1% (14.3-21.7%) | 16.7% (13.3-20.2%) | 15.3% (12.0-18.7%) | 13.9% (10.7-17.4%) | 12.6% (9.5-15.8%) | 11.2% (8.5-14.3%) | 9.8% (6.9-12.6%) | 8.3% (5.9-11.1%) | 7.0% (4.7-9.7%) |
| 3 | 15.5% (12.4-18.6%) | 13.9% (11.2-16.4%) | 12.0% (9.5-14.6%) | 10.4% (8.1-12.8%) | 8.8% (6.7-11.2%) | 7.3% (5.5-9.4%) | 6.0% (4.3-7.7%) | 4.6% (3.2-6.2%) | 3.5% (2.3-5.0%) | 2.6% (1.6-3.7%) |
| 4 | 12.5% (10.3-14.9%) | 10.5% (8.5-12.6%) | 8.6% (6.7-10.6%) | 6.9% (5.3-8.8%) | 5.5% (4.0-7.1%) | 4.2% (2.9-5.5%) | 3.1% (2.1-4.2%) | 2.2% (1.4-3.0%) | 1.4% (0.9-2.2%) | 0.9% (0.5-1.4%) |
| 5 | 9.9% (8.1-12.0%) | 7.8% (6.2-9.6%) | 6.1% (4.7-7.7%) | 4.6% (3.4-6.0%) | 3.3% (2.4-4.5%) | 2.3% (1.6-3.2%) | 1.6% (1.0-2.2%) | 1.0% (0.6-1.5%) | 0.6% (0.3-0.9%) | 0.3% (0.2-0.5%) |

5. World Health Organization. Global tuberculosis report 2022. Geneva, Switzerland: World Health Organization, 2022 19 November 2022. Report No.

6. World Health Organization. WHO consolidated guidelines on tuberculosis. Module 4: treatment-drug-susceptible tuberculosis treatment: World Health Organization; 2022.

7. United Nations Department of Economic and Social Affairs. World Population Prospects 2022 2023. Available from: https://population.un.org/wpp/.

8. Soetaert K, Petzoldt T, Setzer RW. Solving differential equations in R: package deSolve. Journal of statistical software. 2010;33:1-25.

9. World Health Organization. WHO consolidated guidelines on tuberculosis. Module 2: screening-systematic screening for tuberculosis disease: World Health Organization; 2021.

10. World Health Organization. National tuberculosis prevalence surveys: what diagnostic algorithms should be used in future? Geneva, Switzerland: 2023.

11. Kendall EA, Shrestha S, Dowdy DW. The epidemiological importance of subclinical tuberculosis. A critical reappraisal. American Journal of Respiratory and Critical Care Medicine. 2021;203(2):168-74.

12. Zifodya JS, Kreniske JS, Schiller I, Kohli M, Dendukuri N, Schumacher SG, et al. Xpert Ultra versus Xpert MTB/RIF for pulmonary tuberculosis and rifampicin resistance in adults with presumptive pulmonary tuberculosis. Cochrane Database of Systematic Reviews. 2021;(2).

13. Frascella B, Richards AS, Sossen B, Emery JC, Odone A, Law I, et al. Subclinical Tuberculosis Disease—A Review and Analysis of Prevalence Surveys to Inform Definitions, Burden, Associations, and Screening Methodology. Clinical Infectious Diseases. 2021;73(3):e830-e41.

14. Mungai B, Ong 'angò J, Ku CC, Henrion MY, Morton B, Joekes E, et al. Accuracy of computer-aided chest X-ray in community-based tuberculosis screening: Lessons from the 2016 Kenya National Tuberculosis Prevalence Survey. PLOS Global Public Health. 2022;2(11):e0001272.
